# Derivation of four computable 24-hour pediatric sepsis phenotypes to facilitate personalized enrollment in early precise anti-inflammatory clinical trials

**DOI:** 10.1101/2021.12.02.21267016

**Authors:** Yidi Qin, Kate F. Kernan, Zhenjiang Fan, Hyun-Jung Park, Soyeon Kim, Scott W. Canna, John A Kellum, Robert A. Berg, David Wessel, Murray M. Pollack, Kathleen Meert, Mark Hall, Christopher Newth, John C. Lin, Allan Doctor, Tom Shanley, Tim Cornell, Rick E. Harrison, Athena F. Zuppa, Russell Banks, Ron W. Reeder, Richard Holubkov, Daniel A. Notterman, J. Michael Dean, Joseph A. Carcillo, on behalf of the Eunice Kennedy Shriver National Institute of Child Health and Human Development Collaborative Pediatric Critical Care Research Network

**Author notes:** For information regarding this article, Joseph A Carcillo MD, Suite 2000, Faculty Pavilion, UPMC Children’s Hospital of Pittsburgh, 4400 Penn Avenue, Pittsburgh, PA 15421, or.

## Abstract

**Objective:** Thrombotic microangiopathy induced *Thrombocytopenia Associated Multiple Organ Failure* and hyperinflammatory *Macrophage Activation Syndrome* are important causes of late pediatric sepsis mortality that are often missed or have delayed diagnosis. Our objective is to derive computable 24-hour sepsis phenotypes to facilitate enrollment in early precise anti-inflammatory trials targeting mortality from these conditions.

**Design:** Machine learning analysis using consensus k-means clustering.

**Setting:** Nine pediatric intensive care units.

**Patients:** 404 children with severe sepsis.

**Interventions:** 24-hour computable phenotypes derived using 25 bedside variables including C-reactive protein and ferritin.

**Measurements and Main Results:** Four computable phenotypes (PedSep-A, B, C, and D) are derived. Compared to the overall population mean, PedSep-A has the least inflammation (median C-reactive protein 7.3 mg/dL, ferritin 125 ng/mL), younger age, less chronic illness, and more respiratory failure (n = 135; 2% mortality); PedSep-B (median C-reactive protein 13.2 mg/dL, ferritin 225 ng/ mL) has organ failure with intubated respiratory failure, shock, and Glasgow Coma Scale score < 7 (n = 102, 12% mortality); PedSep-C (median C-reactive protein 15.2 mg/dL, ferritin 405 ng/mL) has elevated ferritin, lymphopenia, more shock, more hepatic failure and less respiratory failure (n = 110; mortality 10%); and, PedSep D (median C-reactive protein 13.1 mg/dL ferritin 610 ng/mL), has hyperferritinemic, thrombocytopenic multiple organ failure with more cardiovascular, respiratory, hepatic, renal, hematologic, and neurologic system failures (n = 56, 34% mortality). PedSep-D has highest likelihood of *Thrombocytopenia Associated Multiple Organ Failure* (Adj OR 47.51 95% CI [18.83-136.83], p < 0.0001) and *Macrophage Activation Syndrome* (Adj OR 38.63 95% CI [13.26-137.75], p <0.0001), and an observed survivor interaction with combined methylprednisolone and intravenous immunoglobulin therapies (p < 0.05).

**CONCLUSIONS AND RELEVANCE:** Machine learning identifies four computable phenotypes (www.pedsepsis.pitt.edu). Membership in PedSep-D appears optimal for enrollment in early anti-inflammatory trials targeting *Thrombocytopenia Associated Multiple Organ Failure* and *Macrophage Activation Syndrome*.

**Author’s Comment:** *Question:* Can machine learning methods derive 24-hour computable pediatric sepsis phenotypes that facilitate early identification of patients for enrollment in precise anti-inflammatory therapy trials?

*Findings:* Four distinct phenotypes (PedSep-A, B, C, and D) were derived by assessing 25 bedside clinical variables in 404 children with sepsis. PedSep-D patients had a thrombotic microangiopathy and hyperinflammatory macrophage activation biomarker response, and improved survival odds associated with combined methylprednisolone plus intravenous immunoglobulin therapy.

*Meaning:* Four novel computable 24-hour phenotypes are identifiable (www.pedsepsis.pitt.edu) that could potentially facilitate enrollment in early precise anti-inflammatory trials targeting thrombotic microangiopathy and macrophage activation in pediatric sepsis.

## Introduction

Severe sepsis defined by infection and organ failure contributes to 1 of 5 deaths globally, with over half occurring in children (1). Many of these deaths occur with multiple organ failure (MOF) (2,3) independent from timely shock resuscitation (4,5), implying that dysregulated host immune activation could be targetable in the pediatric intensive care unit (PICU). Among such conditions are immune depression leading to Immunoparalysis associated MOF (IPMOF) (6,7,13,14), thrombotic microangiopathy leading to Thrombocytopenia Associated MOF (TAMOF) (8,9,13,14), and hyperinflammatory Macrophage Activation Syndrome (MAS) driven either by uncontrolled lymphoproliferation manifest as sequential liver failure associated MOF (SMOF) (10,13,14) or by macrophage activation without lymphoproliferation manifest as combined hepatobiliary dysfunction and disseminated intravascular coagulation (11,12,13,14). In the PHENOtyping pediatric sepsis induced Multiple organ failure Study (PHENOMS) (14) we previously reported that these conditions developed at a median of day 3 to 7 of sepsis, with TAMOF and MAS demonstrating 46% mortality, and IPMOF 16% mortality (14). Anti-inflammatory therapies used to reverse TAMOF and MAS include methylprednisolone, intravenous immunoglobulin (IVIG), and plasma exchange (8,15-18).

Our clinical trials challenge is to identify these at-risk children for early enrollment when therapies have their greatest likelihood to succeed. The NIGMS sepsis research working group recommendations call for use of new clinical research approaches in extant clinical data sets (https://loop.nigms.nih.gov/2019/05/recommendations) to characterize septic patients and improve the efficiency of early trials of new sepsis treatments. In this manuscript we use machine learning methods in our extant data set from PHENOMS (14) to derive 24-hour computable sepsis phenotypes based on available bedside clinical variables including C-reactive protein and ferritin (19-21). Our specific goals are to determine 1) if computable 24-hour phenotypes show different risks for development of TAMOF and MAS; and 2) if administration of methylprednisolone, IVIG, and/or plasma exchange by bedside clinicians was associated with survivor interaction in the computable phenotype most likely to develop TAMOF and MAS.

## Materials and Methods Overview

We analyzed blood samples and clinical data obtained from our previously published PHENOMS study (14). Approval was obtained from The University of Utah Institutional Review Board, Central IRB # 70976. Written informed consent was obtained from one or more parents/guardians for each child. Assent was garnered when the child was able. Patients were enrolled from 2015-2017. The CONSORT diagram and details of the clinical study protocol have been previously published (14). Three consented and enrolled children who were excluded from reporting in the parent study manuscript because there was a cap of 81 patients to maximize equalization in enrollment among the centers, are additionally included in this machine learning manuscript. In brief, children qualified for enrollment in PHENOMS if they 1) were between the ages of 44 weeks gestation to 18 years of age; 2) were suspected of having infection meeting two or more of four systemic inflammatory response criteria (22); and 3) had one or more organ failures (23).

DeMerle et al suggests that machine learning phenotypes need to be *clinically relevant, biologically plausible, nonsynonymous, treatment responsive*, and *reproducible* if they are to provide a *‘path forward’* in trial design (24). Our statistical approach is shown in eFigure 1. To derive *nonsynonymous* computable phenotypes, we applied unsupervised clustering methods (25) to clinical and laboratory data available at the first 24 hours of PICU stay with severe sepsis (Table 1, Figure 1, eTables 1-4, eFigures 2-7). To understand *biological plausibility and clinical relevance* we examined correlations between the phenotypes and inflammatory cytokine responses (Figure 2, eTables 5 and 6, eFigure 8); and organ failure and mortality outcomes (Table 2, eTables 7-9, eFigures 9-12). We further assessed development of Immunoparalysis associated MOF (immune depression defined by ex vivo TNF response to endotoxin < 200 pg/ml beyond three days with two or more organ failures) (7,14,26), Thrombocytopenia associated MOF (thrombotic microangiopathy defined by ADAMTS13 activity < 57% of control with platelet count < 100,000/mm^3^ and acute kidney injury with oliguria and serum creatinine > 1 mg/dL) (8,14,26), Sequential Liver Failure associated MOF (lymphoproliferative disease associated with liver failure defined by soluble FAS Ligand > 200 pg/mL with PaO2/FiO2 < 300 and mechanical ventilation followed seven days or later with serum ALT > 100 U/L and bilirubin > 1 mg/dL) (10,14,26) and Macrophage Activation Syndrome (hyperinflammation defined by ferritin > 500 ng/mL with platelet count < 100 K/mm3, INR > 1.5, ALT > 100 U/L and bilirubin > 1 mg/dL) (Table 2, eTable 7, eFig 13) (11,12,14,26).

**Table 1.**
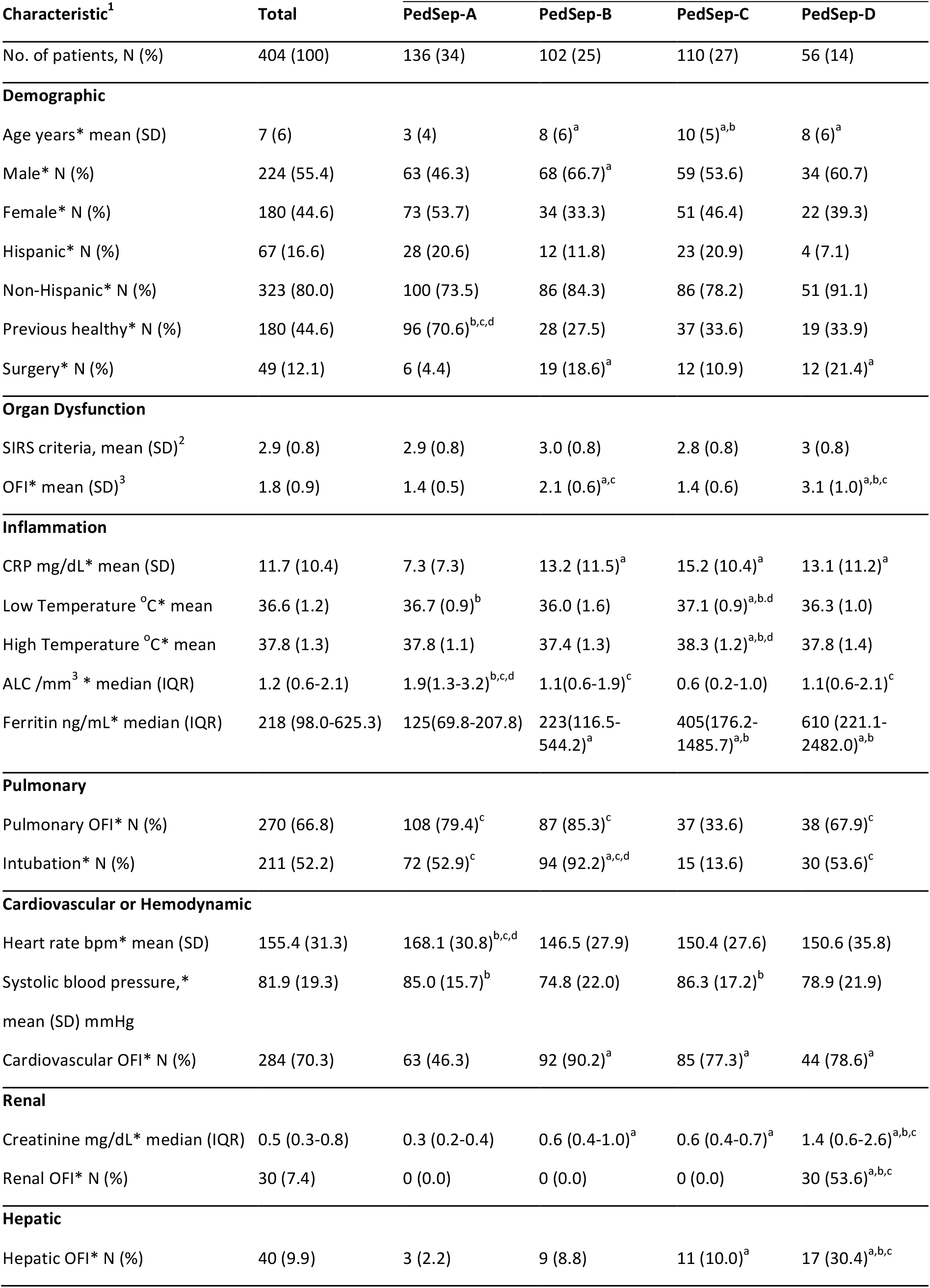
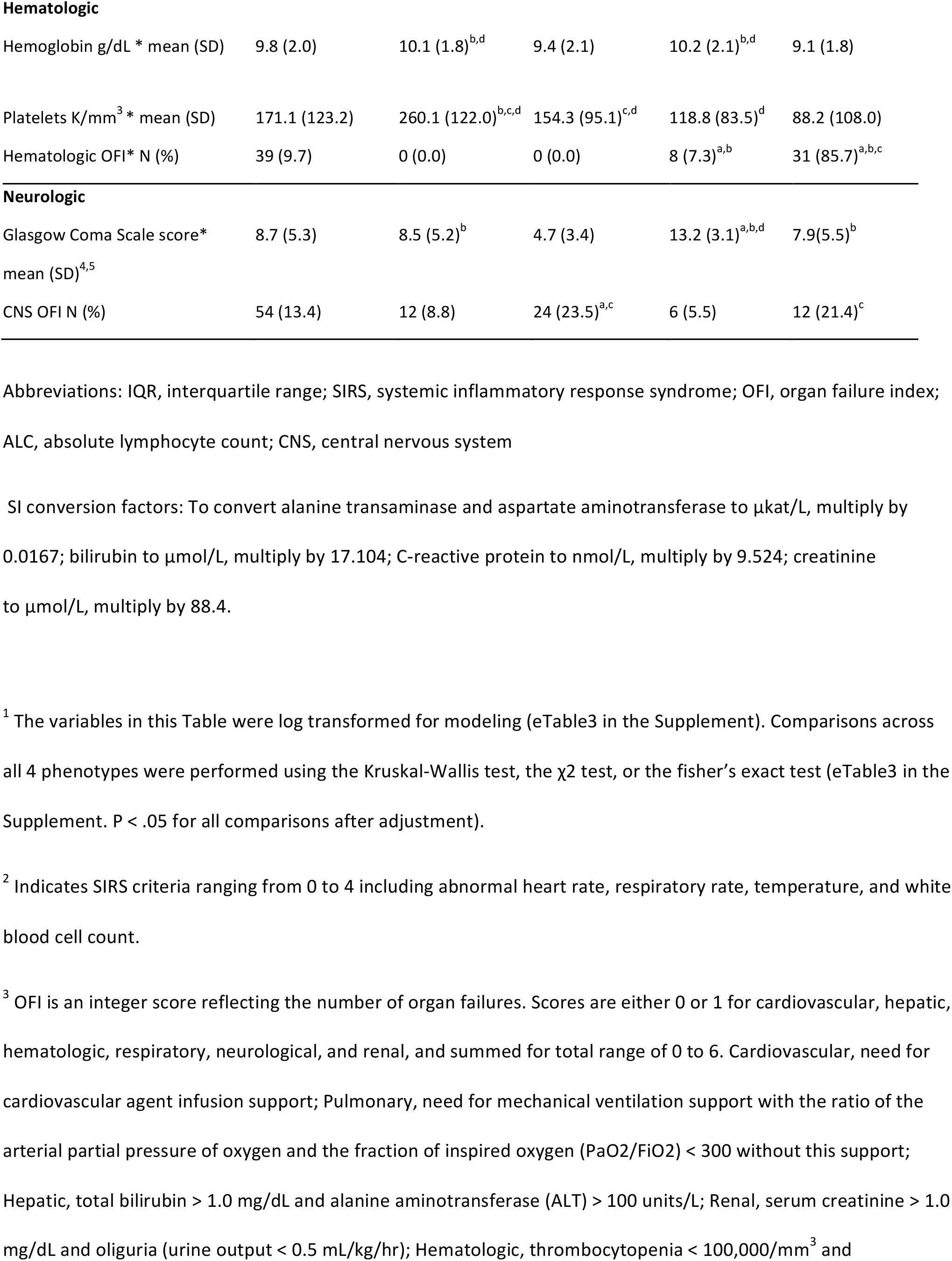
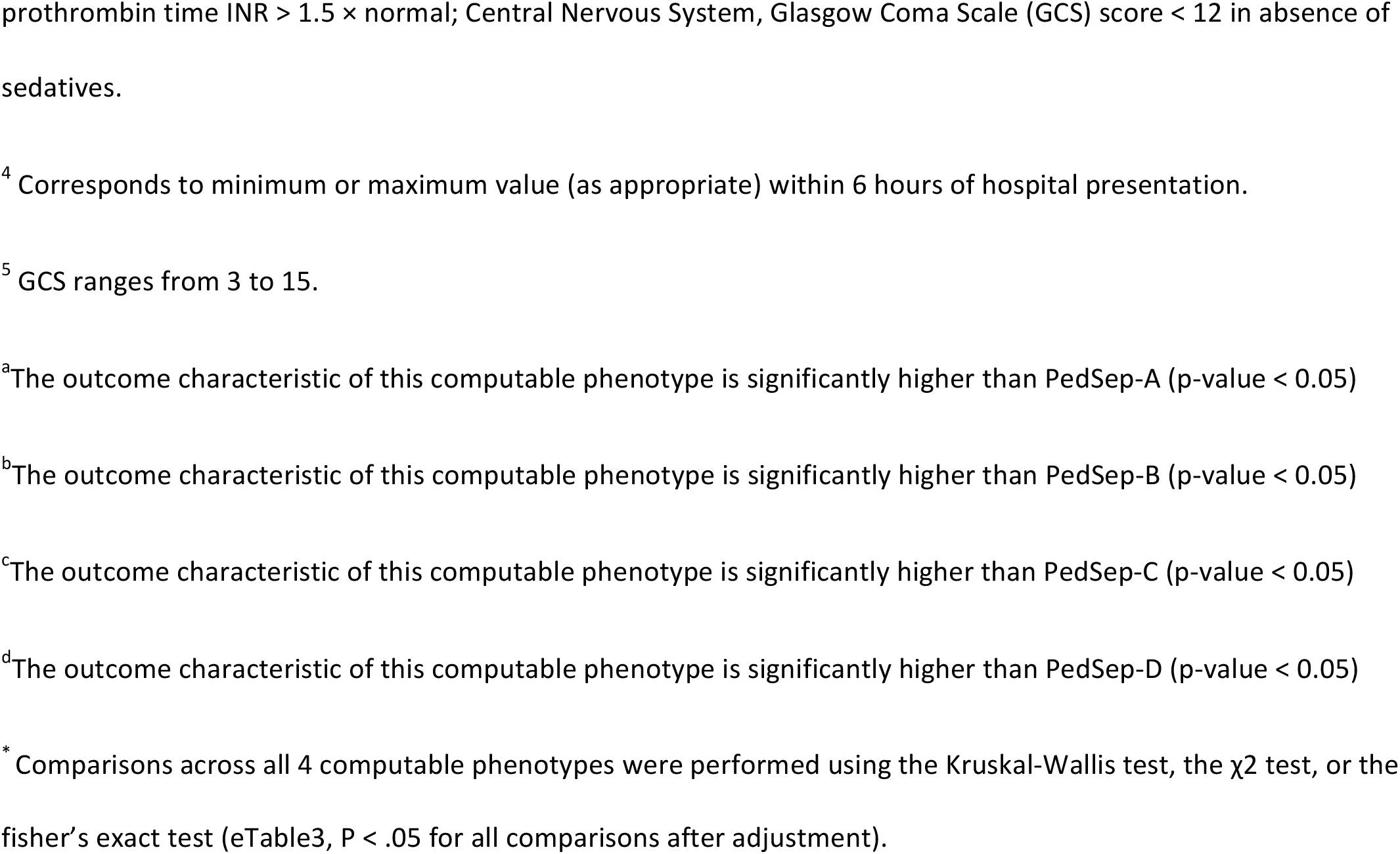
Demographic and Day 1 Clinical Characteristics of the Four Phenotypes.

**Table 2.** Subsequent Outcome Characteristics of the Four Phenotypes.

| Characteristic<sup>e</sup> | Total | PedSep-A | PedSep-B | PedSep-C | PedSep-D |
| --- | --- | --- | --- | --- | --- |
| No. of patients, N (%) | 404 (100) | 136 (34) | 102 (25) | 110 (27) | 56 (14) |
| **Development of Subsequent MOF Empirical Phenotypes** |  |  |  |  |  |
| SMOF, N (%) | 7 (1.7) | 0 (0.0) | 0 (0.0) | 1 (0.9) | 6 (10.7)<sup>a,b,c</sup> |
| TAMOF, N (%) | 37 (9.2) | 0 (0.0) | 6 (5.9)<sup>a</sup> | 3 (2.7) | 28 (50.0)<sup>a,b,c</sup> |
| IPMOF, N (%) | 85 (21.0) | 12 (8.8) | 29 (28.4)<sup>a</sup> | 22 (20) | 22 (39.3)<sup>a</sup> |
| MAS, N (%) | 24 (5.5) | 0 (0.0) | 3 (2.9) | 2 (1.8) | 19 (33.9)<sup>a,b,c</sup> |
| NPMOF, N (%) | 117 (29.0) | 28 (20.6) | 25 (24.5) | 32 (29.1) | 32 (57.1)<sup>a,b,c</sup> |
| **Infections** |  |  |  |  |  |
| Bacterial infection, N (%) | 141 (34.9) | 43 (31.6) | 33 (32.4) | 45 (40.9) | 20 (35.7) |
| Viral infection, N (%) | 114 (28.2) | 60 (44.1)<sup>b,c,d</sup> | 21 (20.6) | 24 (21.8) | 9 (16.1) |
| Fungal infection, N (%) | 4 (1.0) | 0 (0.0) | 1 (1.0) | 0 (0.0) | 3 (5.4) |
| Culture negative, N (%) | 177 (43.8) | 47 (34.6) | 52 (51.0) | 50 (45.5) | 28 (50.0) |
| **Sites of Infections<sup>f</sup>** |  |  |  |  |  |
| Blood, N (%) | 51 (12.6) | 10 (7.4) | 6 (5.9) | 22 (20.0)<sup>a,b</sup> | 13 (23.2)<sup>a,b</sup> |
| Lung, N (%) | 76 (18.8) | 28 (20.6) | 29 (28.4)<sup>a,c,d</sup> | 12 (10.9) | 7 (12.5) |
| Urine, N (%) | 16 (4.0) | 4 (2.9) | 5 (4.9) | 6 (5.5) | 1 (1.8) |
| **Organ Support** |  |  |  |  |  |
| MechVent, N (%) | 366 (90.6) | 134 (98.5)<sup>c</sup> | 101 (99.0)<sup>c</sup> | 79 (71.8) | 52 (92.9)<sup>c</sup> |
| ECMO, N (%) | 30 (7.4) | 5 (3.7) | 9 (8.8) | 6 (5.5) | 10 (17.9)<sup>a</sup> |
| CRRT, N (%) | 52 (12.9) | 1 (0.7) | 7 (6.9) | 7 (6.4) | 37 (66.1)<sup>a,b,c</sup> |
| **Anti-inflammatory Therapies of Interest** |  |  |  |  |  |
| Decadron, N (%) | 94 (23.3) | 50 (36.8)<sup>c,d</sup> | 22 (21.6) | 14 (12.7) | 8 (14.3) |
| Methylprednisone, N (%) | 117 (29.0) | 54 (39.7)<sup>b</sup> | 23 (22.5) | 24 (21.8) | 16 (28.6) |
| IVIg, N (%) | 51 (12.6) | 6 (4.4) | 10 (9.8) | 19 (17.3)<sup>a</sup> | 16 (28.6)<sup>a</sup> |
| Plasma exchange, N (%) | 25 (6.2) | 5 (3.7) | 4 (3.9) | 4 (3.6) | 12 (21.4)<sup>a,b,c</sup> |
| **Outcome** |  |  |  |  |  |
| Length of Stay,<br>median (IQR), d | 9.0 (5.0-17.) | 9.0 (5.8-15)<sup>c</sup> | 10.5 (5.3-17)<sup>c</sup> | 6 (2.3-15) | 12.5 (7-26.5)<sup>c</sup> |
| Mortality, N (%) | 45 (11.1) | 3 (2.2) | 12 (11.7)<sup>a</sup> | 11 (10.0)<sup>a</sup> | 19 (33.9)<sup>a,b,c</sup> |

Abbreviations: SMOF, sequential liver failure associated multiple organ failure; TAMOF, thrombocytopenia associated multiple organ failure; IPMOF, immunoparalysis associated multiple organ failure; MAS, macrophage activation syndrome; NPMOF, new or progressive multiple organ failure; IQR, interquartile range; MechVent, Mechanical Ventilation; ECMO, Extracorporeal Membrane Oxygenation; CRRT, Continuous Renal Replacement Therapies; IVIG, intravenous gamma globulin
<sup>a</sup>The outcome characteristic of this computable phenotype is significantly higher than PedSep-A (p-value < 0.05)
<sup>b</sup>The outcome characteristic of this computable phenotype is significantly higher than PedSep-B (p-value < 0.05)
<sup>c</sup>The outcome characteristic of this computable phenotype is significantly higher than PedSep-C (p-value < 0.05)
<sup>d</sup>The outcome characteristic of this computable phenotype is significantly higher than PedSep-D (p-value < 0.05)
<sup>e</sup>Comparisons across all 4 computable phenotypes were performed using the Kruskal-Wallis test, the $\chi^2$ test, or the fisher's exact test (eTable3, P < .05 for all comparisons after adjustment).
<sup>f</sup>Obtained at the first 3 days

**Figure 1.**
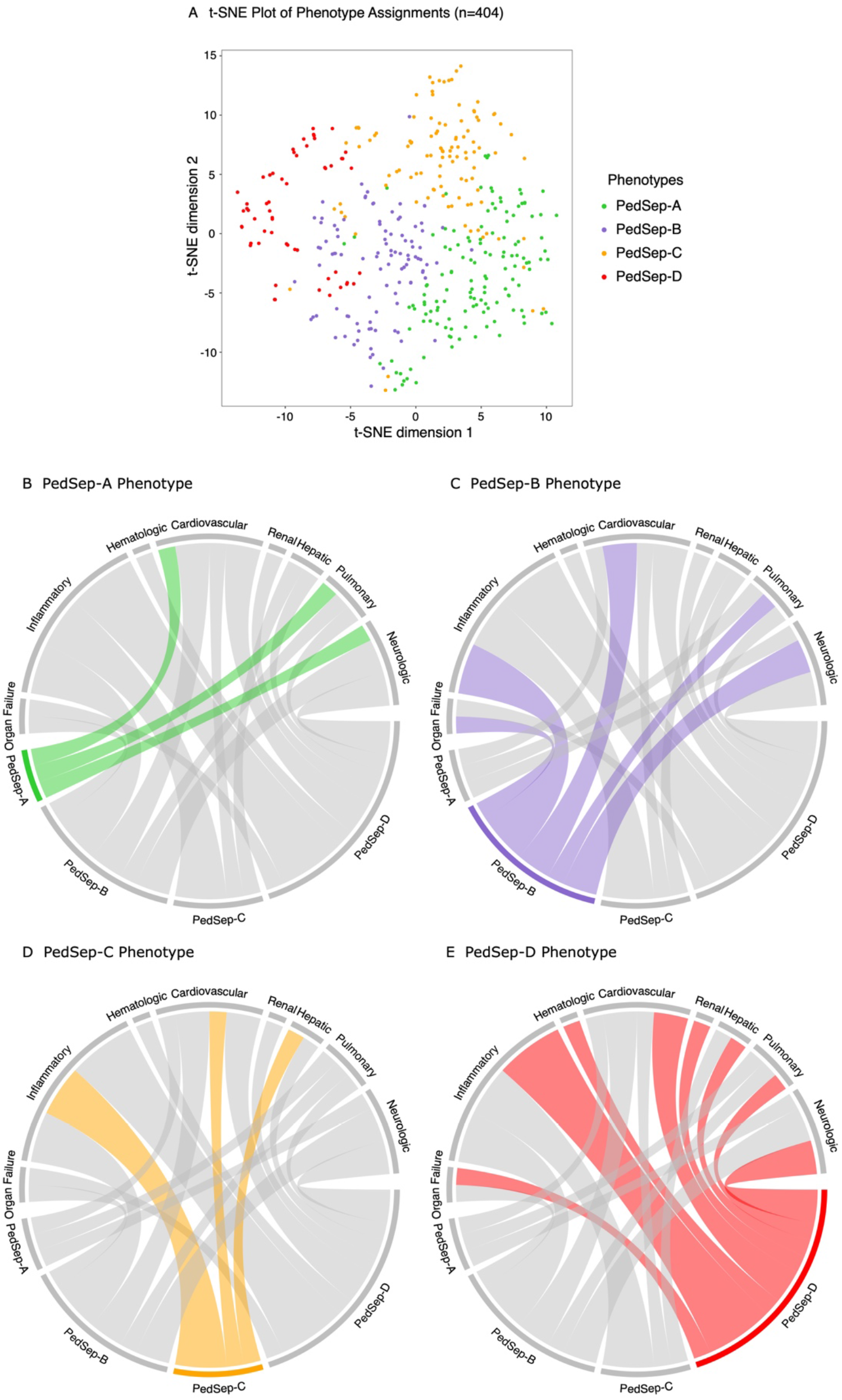
24-hour Phenotype Distribution and Chord plot - In panel A, visualization of phenotypes using t-distributed stochastic neighbor embedding (t-SNE) technique with phenotypes shown in color from the consensus k means clustering analysis visualizes distinction among four phenotypes. In panels B-E, each phenotype is highlighted separately and the ribbons connect to the different patterns of clinical variables and organ system dysfunctions on the top of the circle (Inflammation = Low Temperature, High Temperature, Max CRP, Max Ferritin; Organ Failure = Total OFI; Pulmonary = Pulmonary OFI, Intubation; Cardiovascular = High Heart Rate, Low Systolic Blood Pressure, Cardiovascular OFI; Renal = High Creatinine, Renal OFI; Hepatic = Hepatic OFI; Hematologic = Low Hemoglobin, Low Platelets, Hematologic OFI; Neurologic = Low Glasgow Coma Score Scale, Central Nervous System OFI). The chords connect from an individual phenotype to a category if the group mean involvement of the variables is differs from the overall mean for the entire cohort (see Table 1) specifically lower for Low Temperature, Systolic Blood Pressure, Hemoglobin, Platelets, and Glasgow Coma Scale Score, but higher for all other variables.

**Figure 2.**
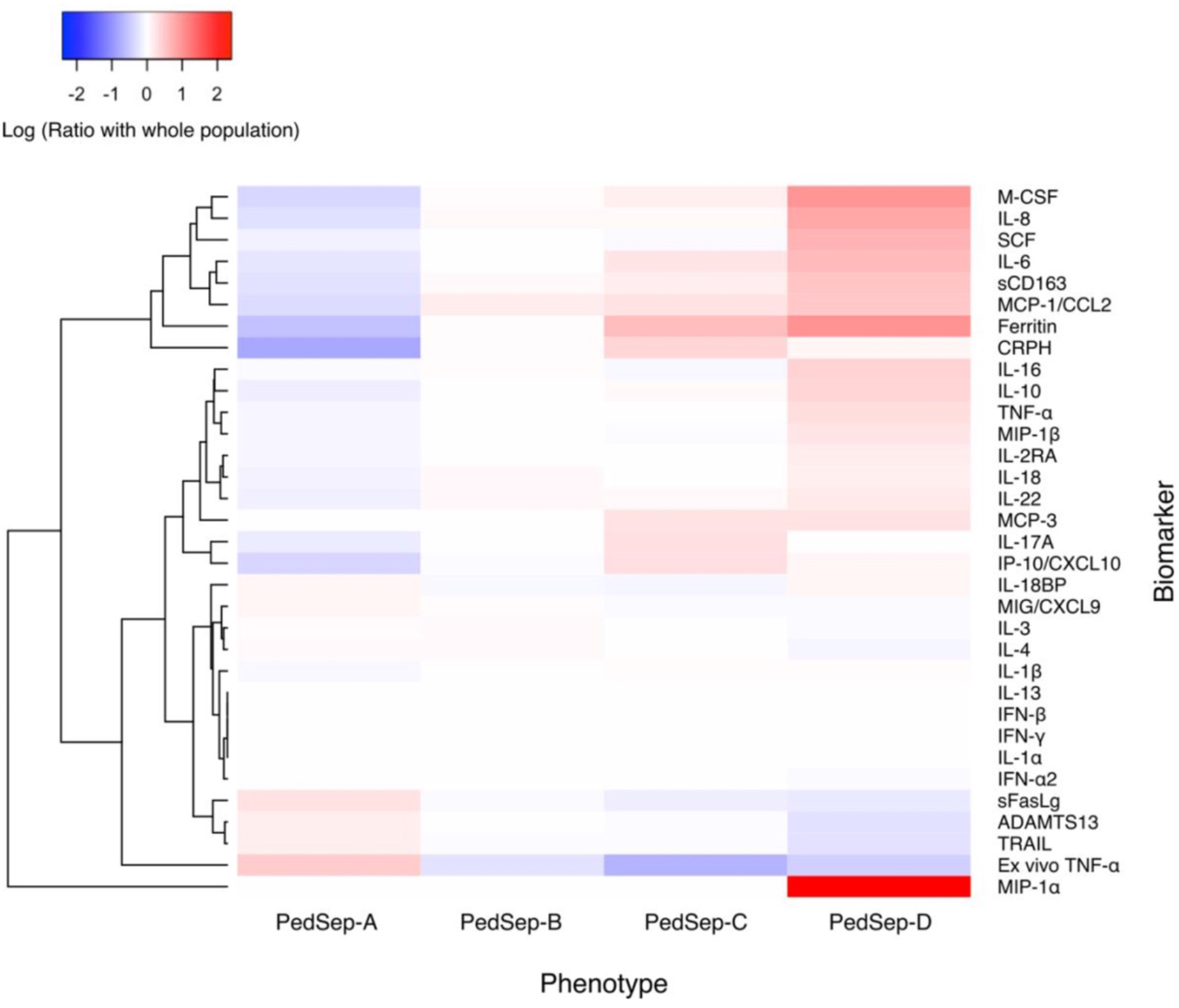
Ratio of inflammatory biomarkers according to 24-hour phenotypes - The cytokine heatmap shows the log ratio of the median biomarker values for various markers of the host response and their hierarchical cluster relationships. Red represents a greater median biomarker value for that phenotype compared with the median for the entire study cohort, whereas blue represents a lower median biomarker value compared with the median for the entire study cohort. For example, M-CSF is lower in PedSep-A than the entire study cohort, and is higher in PedSep-D than the entire study cohort.

In an exploratory analysis of treatment interactions in the phenotypes, we applied elastic net regression analysis to any organ support and anti-inflammatory therapies used by bedside clinicians that were found in univariable analysis to be associated with survival in any of the computable phenotypes or in the population as a whole (p < 0.05) (Figure 3, eTables 10, 11, 12 eFigure 14) (27). Confirmatory logistic regression analysis was performed on anti-inflammatory therapies associated with survivor odds ratio > 10 in the elastic net regression model (eTable 12).

**Figure 3.**
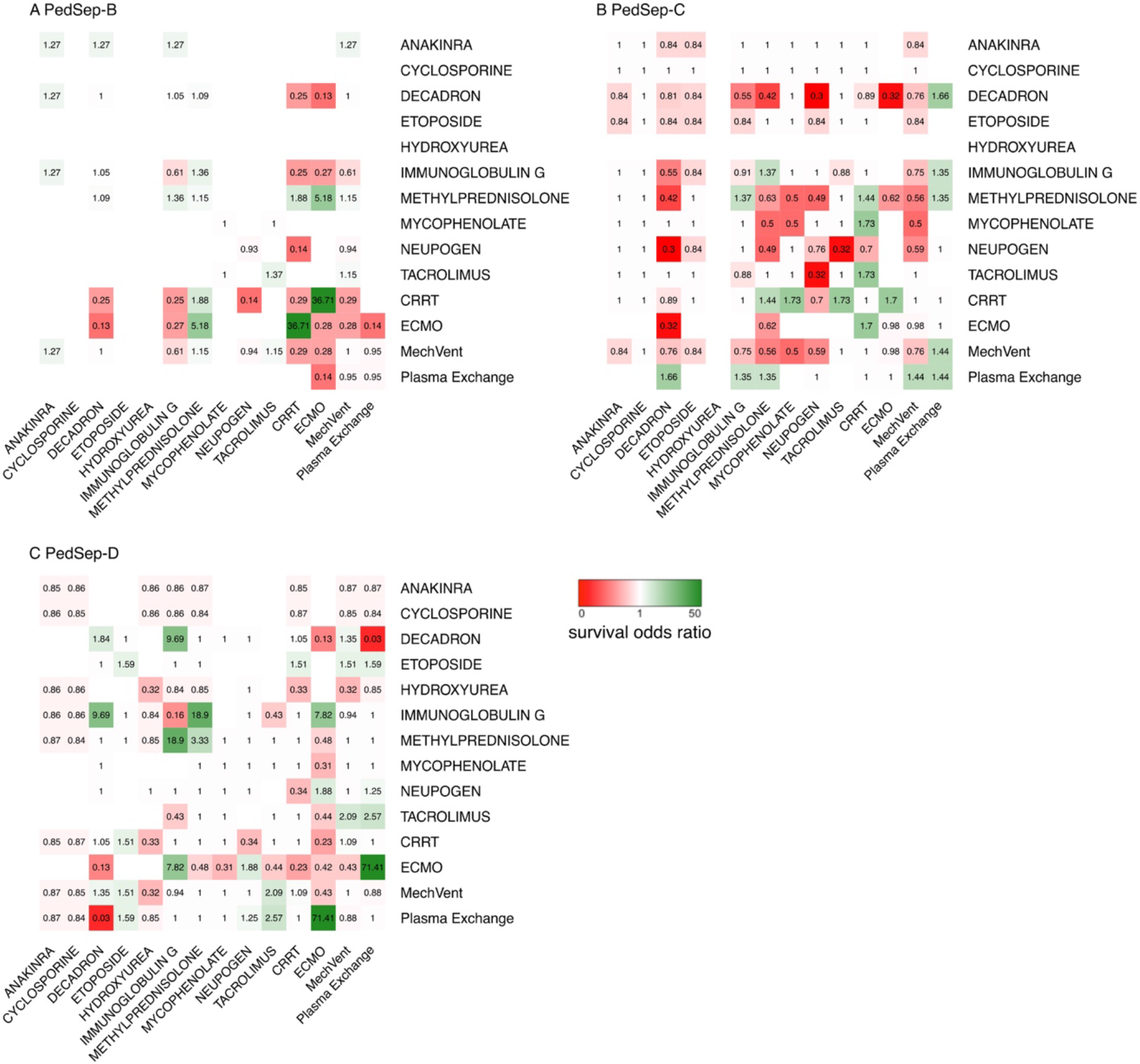
Heterogeneous survival interaction between therapy, and phenotype - Heatmap of Elastic Net Regression analysis shows the association between 14 individual therapies and their 182 combinations (total cells = 196) with survival in PedSep-B, C, and D. The PedSep-A phenotype is not presented due to limited number of deaths. Blank cells have no patients. Values in each cell represent odds ratios of survival, where 1 represents no association with survival. Color in each cell represents direction of effect, where red represents mortality direction, green represents survival direction. Cells located at the diagonal are odds ratio of association from the 14 individual therapies. The other cells represent the survival odds ratio of combinations of these therapies compared to all other combinations. For example, survivors in PedSep-D phenotype are less likely to be treated with IVIG than non-survivors (red); whereas, survivors in PedSep-D are more likely to be treated with combined IVIG + methylprednisolone (green).

### Candidate Clinical Variables for Phenotyping

Of the 52 bedside variables collected in the parent study, only 25 were available at 24 hours with less than 20% missingness and less than 60% correlation with any other variable (Table 1, eTable 1). These included demographic variables (age, gender, ethnicity, previous health status, post-op status), PRISM related vital signs and laboratory values (systolic blood pressure, heart rate, Glasgow Coma Scale score, hemoglobin, creatinine, platelet count, intubation status), markers of inflammation (temperature, number of SIRS criteria, lymphocyte count, C-reactive protein level, ferritin level), and organ failures (Central Nervous System = Glasgow Coma Scale < 12 not explained by use of sedation; Cardiovascular = Requirement for vasoactive agents for Systolic Blood Pressure < 5th percentile for age; Respiratory = PaO2/FiO2 ratio < 300 requiring mechanical ventilation; Renal = oliguria and serum creatinine > 1 mg/dL; Hepatic = ALT > 100 and Bilirubin > 1 mg / dL; Hematologic = Platelet Count < 100K and INR > 1.5) (10,14,26). For each PRISM variable we extracted the most abnormal value in the first 6 hours. For each inflammation and organ failure variable we extracted the most abnormal value within 24 hours.

### Biological Correlates and Outcomes

We studied 33 biomarkers including 31 cytokines and two functional assays; whole blood *ex vivo* TNF response to endotoxin as a marker of immune depression (6,7,14,26), and ADAMTS 13 activity as a marker of microvascular thrombosis in the presence of thrombocytopenia (6,14,26). Plasma for cytokine measurement was divided into three assays. IL-18, IL-18BP, and CXCL9 were measured at 25-fold dilution (28). IFN *α*, sCD163, and IL-22 were measured by Bioplex inflammatory flex-set assay per manufacturer’s instructions (Bio-Rad). The remainder were measured by Bioplex Group I/II flex-set assay (Bio-Rad). All cytokines were measured on a BioPlex 200 System (Bio-Rad). The functional assays were measured as previously described (6,7,8,14,26).

The primary outcome was hospital mortality. Secondary outcomes included development of new or progressive MOF defined as development of new organ failure(s) after day one (2); length of stay in the PICU; subsequent development of Immunoparalysis (7,8,14,26), Thrombocytopenia associated MOF (8,14,26), Sequential liver failure associated MOF (10,14,26), and Macrophage Activation Syndrome (11,12,14,26); as well as use of mechanical ventilation, and extracorporeal therapies.

Adjusted odds ratios controlling for age, sex, ethnicity, race, and total PRISM score, were calculated for primary and selected secondary outcomes. For summary analyses the threshold for statistical significance was less than 0.05 for two - sided tests after adjustment for multiple testing. All analyses were performed with R version 3.6.2.

## Results

### Derivation of Clinical Sepsis Phenotypes

The consensus k-means clustering models were used because the method provides *nonsynonomous* agnostic clusters and has a 1,000 iterations step to assure internal consistency (19). We found a 4-class model was the optimal fit, with phenotypes we named PedSep-A, B, C and D (eFigure 4). Consensus matrix plots and the relative change under cumulative distribution function curve implied little statistical gain by increasing to a 5 or 6 class model, with penalty of overfitting. The size and characteristics of the 4-class model appear in Table 1 and Figure 1. They ranged in size (from 14% to 34% of the cohort) and differed in clinical characteristics and organ dysfunction patterns (Table 1, eTable 3, Figure 1, eFigures 5 and 9). With the exception of the SIRS criteria number, the other 24 variables differed among the phenotypes. Compared to all other phenotypes PedSep-A patients were younger and previously healthy, with the lowest CRP and ferritin levels, the highest lymphocyte and platelet counts, highest heart rate, and lowest creatinine; PedSep-B patients were most likely to be intubated and had the lowest Glasgow Coma Scale Score; PedSep C patients had the highest temperature and Glasgow Coma Scale score, least pulmonary failure, and lowest lymphocyte count; and PedSep-D patients had the highest creatinine and number of organ failures, including renal, hepatic, and hematologic organ failure, with the lowest platelet count. On average, PedSep-B and D patients had multiple organ failure whereas PedSep-A and C patients did not. Ferritin levels were highest in PedSep-C and PedSep-D distinguishing them from PedSep-A and B (Table 1, eTable 3, eFigures 5 and 9)

### Correlation of Phenotypes with Biomarker Profiles

The inflammatory biomarker profiles differed across the four computable phenotypes. Inflammation (as evidenced by cytokine signature) increased and immune response (whole blood *ex vivo* TNF response to endotoxin) and coagulation function (ADAMTS13 activity) decreased going across PedSep-A, B, C, and D (Tables 5 and 6, Figure 2, eFigure 8). PedSep-A showed the least inflammation with the lowest M-CSF, IL-8, IL-6, sCD163, MCP1/CCL2, ferritin, C-reactive protein, IL-10, IL-22, and MIP 1*α* levels overall; lower IL-17a and IP10/CXCL10 than PedSep-C; and lower IL-18 and IL2Ra than PedSep-D. PedSep-A had the best immune and coagulation function with normal whole blood ex vivo TNF response to endotoxin (> 200 pg/mL) and ADAMTS 13 activity. In contrast PedSep-D had the most profound inflammatory response with highest M-CSF, IL-8, SCF, sCD163, IL-16, IL-10, TNF, and MIP1*α* levels; and thrombotic microangiopathic response with lowest ADAMTS13 activity decreased to < 57% of control with thrombocytopenia. Consistent with this increased inflammation response the macrophage inhibitor TRAIL was reduced in PedSep-D compared to PedSep-C.

### Relationship with Infection, Organ Support Needs, and Hospital Mortality

PedSep-A had more viral infections, PedSep-B had more pneumonia, and PedSep-C and D had more blood infections. Patients in PedSep-C had the least mechanical ventilation and the shortest length of stay. Patients in PedSep-D required more extracorporeal membrane oxygenation than in PedSep-A, and the most continuous renal replacement therapy (CRRT) overall. PedSep-A patients required the least CRRT (Table 2, eTable 7).

Hospital mortality was 2% in PedSep-A, 12% in PedSep-B, 10% in PedSep-C, and 34% in PedSep-D (PedSep B vs A Adj OR 4.11 95% CI [1.11-19.96] p = 0.048; PedSep C vs A Adj OR 4.35 95% CI [1.23-20.43] p = 0.034; PedSep D vs A Adj OR 17.25 95% CI [4.93-92.06] p = 4.42E-05; PedSep D vs B Adj OR 4.20 95%CI [1.84-9.97] p = 0.0008; and PedSep D vs C Adj OR 3.97 95% CI 1.62-10.14] p = 0.003) (Table 2, eTable 7).

The mortality curves show all deaths in PedSep-A occurred before seven days; whereas, deaths in PedSep-B, C, and D continued to accrue after seven days (eFigure 10). Mortality was associated with Glasgow Coma Scale score < 12, decreased TNF and IL-2Ra levels, and increased MCP3 levels in PedSep-A; increased IL-6, IL-8, and MCP1/CCL2 levels in PedSep-B; high ferritin, lymphopenia, lower temperature, higher blood pressure, and increased IL-8 levels in PedSep-C; and, hyperferritinemia, chronic illness, increased MIP-1*α*, IL-8, and IL-10 levels, and decreased IL-18 and sFASL levels in PedSep-D (eTables 8 and 9, eFigure 11 and 12).

### Relationship with Development of Immunoparalysis, TAMOF, SMOF, and MAS

On average, children in PedSep-A and PedSep-C developed less than two organ failures; children in PedSep-B developed more than two organ failures; and children in PedSep-D developed more than three organ failures over 28 days (eFigure 10). Children in PedSep-D had the highest proclivity to develop Immunoparalysis (Adj OR 2.40 95% CI [1.25-4.53; p = 7.20E-03), new and progressive organ failure (Adj OR 4.03 95% CI [2.19-7.55]; p = 9.48E-06), Thrombocytopenia associated MOF (Adj OR 47.51 95% CI [18.83-136.83]; p = 1.25E-14), Sequential liver failure associated MOF (Adj OR 61.56 95% CI [8.93-1,282.58]; p = 3.80E-04), and Macrophage Activation Syndrome (Adj OR 38.63 95% CI [13.26-137.75]; p = 4.61E-10). Immunoparalysis and Thrombocytopenia associated MOF also occurred more commonly in children in PedSep-B and D compared to those in PedSep-A (Table 2, eFigure 13).

### Heterogeneous treatment interactions with use of anti-inflammatory therapies

All 3 organ support therapies, and 11 of 41 anti-inflammatory therapies were associated with outcome in univariable analysis (eTable 10 and 11) and included in the exploratory elastic net regression analysis (Figure 3, eFig 14) (27). This was not performed in PedSep-A because mortality was very low at 2%. The constructed elastic net regression heatmaps visualize heterogeneous survival association patterns across PedSep-B, C, and D (Figure 3). Survivor odds ratios > 10-fold with use of anti-inflammatory agents were only observed in PedSep-D, in those patients receiving combined corticosteroids plus IVIG, and in extracorporeal membrane oxygenator patients receiving plasma exchange.

Logistic regression analysis confirmed a survivor interaction in PedSep D patients receiving methylprednisolone plus IVIG compared to either methylprednisone or IVIG alone (Methylprednisolone Adj OR 0.50 95% CI [0.073-3.75] p = 0.47; IVIG Adj OR 0.16 95% CI [0.018-1.02] p = 0.07; Methylprednisolone + IVIG Adj OR = 35.13; 95% CI [1.57-722.68] p = 0.042); and also a survivor interaction with use of combined methylprednisone + IVIG in PedSep D patients compared to PedSep B + C patients (PedSep-D vs B + C Adj OR = 0.19 95% CI [0.09-0.41] p < 0.001; Methylprednisolone + IVIG combination Adj OR = 0.22 95% CI [0.06-0.89] p = 0.022; PedSep-D * Methylprednisolone + IVIG combination Adj OR = 16.67 95% CI [1.64-500] p = 0.029) indicating that the survival outcome interaction associated with this combination therapy is specific to PedSep-D (eFig 14). Logistic regression analysis was not statistically significant for combined ECMO and plasma exchange therapy (eTable 12).

## Discussion

Machine learning analysis of our extant PHENOMS dataset identifies four computable 24-hour phenotypes meeting three of five ‘path forward’ criteria (24) providing impetus for further evaluation in new pediatric sepsis studies. The phenotypes demonstrated *clinical relevance* with differences in types of infections, organ failures, need for organ support therapies, outcomes, and proclivity to development of TAMOF and MAS. Consensus k-means clustering and t-SNE analyses demonstrated that the phenotypes are *nonsynonymous*. The differences in cytokine profiles provide *biological plausibility* for these phenotypes having different inflammation responses, highlighted in PedSep-D by decreased ADAMTS13 with TAMOF and increased MIP 1*α* with MAS. Exploratory modeling of treatment interactions with survival using elastic net regression showed a signal with combined methylprednisone plus IVIG therapy in PedSep D; however, wide confidence intervals in the confirmatory logistic regression analysis provides impetus for further evaluation in new studies. We are presently assessing *treatment responsiveness* and *reproducibility* in the ongoing 500 patient *Second Argentinian Pediatric Sepsis Epidemiology Study* (PI Roberto Jabornisky), and in the placebo arm of our NICHD network’s ongoing 1,000 patient *Personalized Immunomodulation in Pediatric Sepsis and Multiple Organ Dysfunction trial*.

PedSep-A is characterized by younger previously healthy children with respiratory failure and the least increased inflammation. This resembles the adult *α* phenotype in the SENECA trial (19), and also the MARS 3 and sepsis response signature 2 endotypes, which found predominant expression of adaptive immune and B-cell developmental pathways (29-31). Mortality in PedSep-A was low at 2% and did not increase after 7 days, making anti-inflammatory clinical trials directed to survival less feasible.

PedSep-B is characterized by multiple organ failure requiring intubation for more severe respiratory failure, shock, and central nervous system dysfunction with increased C-reactive protein levels and 12% mortality. This is reminiscent of children reported in the *Life After Pediatric Sepsis Evaluation* study (32); the shock with hypoxia phenotype in adult sepsis induced MOF (33); and the severe hypoxia, altered mental status, and shock phenotype in pediatric MOF (34).

PedSep-C is distinguished by cardiovascular failure and relative absence of need for intubation, in the presence of elevated C-reactive protein, high ferritin, and lymphopenia, with 10% mortality. This is reminiscent of the Toxic Shock (TSS) - Kawasaki syndrome phenotype currently being considered as PMIS/MIS-C syndrome (35-39). Similar to TSS and Kawasaki’s, our PedSep-C patients showed elevated IL-17a and IP10/CXCL10 levels (40-42).

PedSep-D patients had cardiovascular, respiratory, liver, renal, hematologic, and neurologic dysfunction with 34% mortality; clinical features shared by the adult *δ* phenotype characterized in the SENECA study using electronic health record criteria for Sepsis-3 (19); the shock with thrombocytopenia pediatric MOF phenotype (34); and previously reported subclasses including the hyperinflammatory sub-phenotype reported in acute respiratory distress syndrome, a condition commonly related to sepsis (43,44). It also resembles sepsis endotypes derived using transcriptomic analyses of circulating immune cells, specifically the inflammopathic cluster known as sepsis signature 1, or the Molecular Diagnosis and Risk Stratification of Sepsis [MARS] 2 cluster (29-31).

PedSep-D is specifically characterized by hyperferritinemic (ferritin > 500 ng/mL), thrombocytopenic (platelet count < 100 K) multiple organ failure with the highest likelihood of new and progressive multiple organ failure, accruing mortality after 7 days. PedSep-D membership identifies children with highest proclivity for decreased ADAMTS 13 activity with Thrombocytopenia Associated MOF, and increased MIP 1*α* with Macrophage Activation Syndrome. Methylprednisone and IVIG therapy have been associated with survival in hyperferritinemic, thrombocytopenic sepsis and macrophage activation syndrome (15,16), and were reminiscently associated with a survivor interaction in PedSep-D.

Although the PHENOMS database is the largest available longitudinal multiple center pediatric sepsis induced MOF study with CRP and ferritin levels (14) it brings inherent limitations this machine learning analysis. Definitions of sepsis and organ failure are limited to those used in this parent study. Only 25 out of 52 available clinical and laboratory variables had < 20% missingness without covariance for inclusion in the derivation. Only 33 additional biomarkers (7,8,10,26,45) were performed for assessment of biological plausibility. Survival interactions could only be assessed for the therapies given by bedside clinicians as part of their standard care of patients. Reproducibility cannot be addressed in one study.

### Conclusions

Machine learning analysis of the PHENOMS database identifies four novel computable 24-hour pediatric sepsis phenotypes and provides a computable tool allowing clinical researchers to perform bedside identification of individual patient phenotype (www.pedsepsis.pitt.edu). Among these, PedSep-D membership identifies children most likely to benefit from early enrollment in anti-inflammatory trials targeting Thrombocytopenia Associated MOF and Macrophage Activation Syndrome. Over the next five years, reproducibility will be assessed in two ongoing pediatric sepsis studies to determine whether this computable tool can be used in the future to improve the efficiency of early trials of new sepsis treatments targeting thrombotic microangiopathy and macrophage activation in children.

## Supporting information

eSupplement

## Data Availability

All data produced in the present work are contained in the manuscript

https://www.pedsepsis.pitt.edu

## ACKNOWLEDGMENTS

Clinical Research Investigation and Systems Modeling of Acute illness center: Ali Smith, BS; Octavia Palmer, MD; Vanessa Jackson, AA; Renee Anderko, BS, MS. Children’s Hospital of Pittsburgh: Jennifer Jones, RN; Luther Springs. Children’s Hospital of Philadelphia: Carolanne Twelves, RN, BSN, CCRC; Mary Ann Diliberto, BS, RN, CCRC; Martha Sisko, BSN, RN, CCRC, MS; Pamela Diehl, BSN, RN; Janice Prodell, RN, BSN, CCRC; Jenny Bush, RNC, BSN; Kathryn Graham, BA; Kerry Costlow, BS; Sara Sanchez. Children’s National Hospital: Elyse Tomanio, BSN, RN; Diane Hession, MSN, RN; Katherine Burke, BS. Children’s Hospital of Michigan, Central Michigan University: Ann Pawluszka, RN, BSN; Melanie Lulic, BS. Nationwide Children’s Hospital: Lisa Steele, RN, CCRC; Andrew R. Yates, MD; Josey Hensley, RN; Janet Cihla, RN; Jill Popelka, RN; Lisa Hanson-Huber, BS. Children’s Hospital Los Angeles and Mattel Children’s Hospital: Jeni Kwok, JD; Amy Yamakawa, BS. Children’s Hospital of Washington University of Saint Louis: Michelle Eaton, RN. Mott Children’s Hospital: Frank Moler, MD; Chaandini Jayachandran, MS, CCRP. University of Utah Data Coordinating Center: Teresa Liu, MPH, CCRP; Jeri Burr, MS, RN-BC, CCRC, FACRP; Missy Ringwood, BS, CMC; Nael Abdelsamad, MD, CCRC; Whit Coleman, MSRA, BSN, RN, CCRC.

