## Supplementary material for "Derivation of four computable 24-hour pediatric sepsis phenotypes to facilitate personalized enrollment in early precise anti-inflammatory clinical trials": eSupplement: Qin Kernan Medrxiv Supplements.pdf

#### Supplementary Digital Content Detailed Overview of Statistical Methods

##### A. Variable selection and missing data imputation for input in Consensus k-means clustering

The workflow of our statistical methods is presented in eFigure 1. The parent study collected 6 demographic variables and 46 clinical variables (eTable 1). For clustering, we selected variables based on correlation, distribution, and missingness (eTable 2, eFigure 1). First, highly correlated variables (Pearson correlation > 0.6) were removed. Among the remaining 41 variables, a total of 25 variables had missingness < 20% and were selected and scaled for computable phenotype determination. Log transformation was used for non-parametric variables.

Further, to impute the missing data in the 25 selected variables, multiple imputation with chained equation (MICE) was used (1). MICE is based on the Fully Conditional Specification (FCS) approach, where each incomplete variable is imputed by a separate model. Using FCS, MICE can impute mixes of continuous, binary, unordered categorical and ordered categorical data. In running MICE, we assumed that missing data is conditional on observed data and is following the pattern of "missing at random".

##### B. Consensus k-means clustering derivation of ***nonsynonomous*** cluster phenotypes

Consensus k-means clustering was used to identify the phenotype membership of patients based on different number of clusters (k) as performed in the SENECA study. We then identified the optimal number of phenotypes (clusters) according to diagnostic plots.

To determine the membership of each patient, consensus clustering takes advantage of subsampling techniques so that perturbations of the original data can be simulated. In each subsampling run, the k-means algorithm was applied to the perturbed data sets for a given k. With this setting, the method calculates a "consensus value" for each pair of patients. This consensus value can be interpreted as the frequency the two patients are assigned to the same phenotype. It ranges from 0 to 1, with larger values indicating stronger consensus. Then, a hierarchical clustering is applied on consensus value matrix to obtain final cluster assignment for each subject.

To determine the optimal number of clusters, we assessed a combination of phenotype size, separation of the consensus matrix heatmaps, characteristics of the consensus cumulative distribution function plots, and adequate pairwise–consensus values between cluster members (eFigure 4). The consensus matrix heatmap is a plot having patients as both rows and columns. The consensus value is the frequency the two patients are assigned to the same phenotype among 1000 iterations. It ranges from 0 (white, interpreted as two patients are never clustered together) to 1 (dark blue interpreted as two patients are always clustered together). A clear separation of white and dark blue blocks in the heatmap is an indicator of good partitioning. The Consensus CDF plot shows the cumulative distribution functions of the consensus matrix for each k, estimated by a histogram of 100 bins. It is used to determine at what number of clusters the CDF reaches an approximate maximum; thus, consensus and cluster confidence is at a maximum at this k. It is usually used together with the Delta area plot to determine the optimal k. Usually, an "elbow" in the Delta area plot is an indicator of the optimal k. The Cluster-consensus plot shows the cluster-consensus value of clusters at each k. This is the mean of all pairwise consensus values between a cluster's members. High values indicate a cluster has high stability and low values indicate a cluster has low stability. We used 0.5 as a cut off for diagnostic purposes.

Our choice of consensus k-means clustering was based on the following two reasons. Firstly, consensus clustering provides a solution to represent the most common assignment across multiple runs of a clustering algorithm. Therefore, it is able to provide qualitative and quantitative measurements for internal validation purposes, which contributes to determining the number and the stability of the discovered clusters. Secondly, we chose to use k-means as the inner-loop clustering algorithm based on the data structure indicated by the OPTICS plot (eFigure3) (2). The OPTICS plot shows a smooth rise instead of clearly partitioned valleys, which implies that the data cannot be arranged into discrete natural groupings. In this case, a partitioning clustering method, such as k-means, is more appropriate than hierarchical clustering methods.

##### C. Comparison of Latent class analysis derived groups to Consensus k means phenotypes

To assess and confirm underlying phenotype structure and stability of individual assignment of the four phenotypes we applied a second method namely latent class analysis (LCA) (3) to the same dataset (eTable 4, eFigure 5-7) as was done in the SENECA study. After running LCA with different numbers of clusters, we separately used Bayesian information criteria (BIC), group size, membership of patients and clinical characteristics of phenotype groups (eTable 4) to confirm that four underlying phenotypes is optimal using LCA in our data (eFigure 6). We further visualize and find similar side by side Rank of variable contributions based on the LCA as with the consensus k means method (eFigure 5), and visualize reasonable patient membership overlap between consensus k means and LCA in an alluvial diagram (eFigure 7).

**D. Dissimilarity visualization of Consensus k means phenotypes**

In order to assess dissimilarity among the four consensus k means phenotypes we used 1) a t-distributed stochastic neighbor embedding (t-SNE) plot labeled by outcomes of interest (Figure 1); 2) chord diagrams in terms of clinical characteristics and organ dysfunction patterns (Figure 1); and 3) variable contributions to pairwise phenotype discrimination (eFigure 5).

**E. Heterogeneity of biomarkers and primary outcomes among phenotypes**

Following the consensus k means phenotype determination, we correlated the identified phenotypes with their biomarkers (Figure 2, eTable 5 and 6, eFigure 8) and investigated their relationships to mortality (eTable 7-9, eFigures 10-12), organ failure (eFigures 9 and 10), and MOF pathobiology groups (Table 2, eFigure 13). To determine the correlation of the 24-hour phenotypes with 33 biomarkers of the host response (Figure 2, eTable 5 and 6, eFigure 8) we compared mean and standard deviation, median and interquartile range (IQR) for continuous data, and the ratio of the cases in binary data. To investigate the relationship of the phenotypes to primary outcomes (mortality, MOF groups), we estimated the association between phenotype and outcomes with a multivariate model adjusting for demographic variables (age, sex, race, ethnicity) and PRISM score. We also generated phenotype specific curve plots to assess differences in mortality and number of organ failures over time (eFigure 10).

**F. Exploratory analysis of heterogeneity of treatment interactions with phenotypes**

Within each of the identified phenotypes, we first evaluated which of 41 candidate anti-inflammatory and 3 organ support therapies given in the parent study by bedside clinicians were associated with survival in univariate logistic regression (eTable 10, 11). For the 14 significant individual treatments, we further applied Elastic Net regression to investigate their interactive effects on survival within each of the phenotypes (Figure 3, eFig 14).[26] Two treatment combinations in PedSep-D were found by elastic net regression analysis (corticosteroids plus IVIG in overall patients; and added plasma exchange in ECMO patients) to have a survival Odds Ratio effect > 10. We performed traditional multivariable logistic regression to validate interaction between these therapies in PedSep-D patients (eTable 12).

**G. Other information**

For summary analysis, we presented continuous data as mean (SD) or median (IQR) and categorical data as count number (%). For comparison, we used Kruskal-Wallis tests for continuous data and the chi square test for categorical data. Fisher exact tests were applied for cells containing less than 5 samples. The threshold for statistical significance was less than 0.05 for two-sided tests after adjustment for multiple testing. Holm–Bonferroni correction was applied to correct for multiple testing. Analyses were performed with R version 3.6.2.

**H. Computable prediction tool of individual membership in phenotypes**

We also developed a computable tool (www.pittsepsis.edu) that allows one to categorize the phenotype of a new individual patient into one of the four phenotypes at the bedside. Specifically, we first used the same pipeline to standardize and normalize the 25 input variables of the new patient as described above. Then we calculated the Euclidean distance from this patient to the centroid of each phenotype derived from the unsupervised consensus k-means clustering. Comparing the distances to four phenotype centroids, we assign the patient to the phenotype with the shortest distance.

1. Newgard CD, Haukoos JS. Advanced statistics: missing data in clinical research—part 2: multiple imputation. Acad Emerg Med. 2007;14(7):669-678.
2. Ankerst M. OPTICS: ordering points to identify the clustering structure. SIGMOD Rec. 1999;28(2): 49-60. doi:10.1145/304181.304187.
3. Rindskopf D, Rindskopf W. The value of latent class analysis in medical diagnosis. Stat Med. 1986;5 (1):21-27. doi:10.1002/sim.4780050105.

#### Supplementary Digital Content eTables 1-12 and eFigures 1-14

eTable 1. List of all candidate variables

eTable 2. Missing data (no., %) for demographic and day 1 clinical variables

eTable 3. Statistical comparison of demographic and day 1 clinical characteristics (n = 404)

eTable 4. Statistical output from latent class analysis

eTable 5. Biomarker levels at Day 1 in PedSep A, B, C, and D

eTable 6. Statistical comparison of biomarker levels measured at day 1 in PedSep-A, B, C, and D

eTable 7. Statistical Test Results of Subsequent Outcome Characteristics (n = 404)

eTable 8. Statistical test results of association between day 1 characteristics and mortality in PedSep-A to D

eTable 9. Statistical test results of association between biomarkers and mortality in PedSep A, B, C, and D

eTable 10. Statistical test result of 44 candidate therapies association with outcome

eTable 11. Median duration of common anti-inflammatory/immune medications

eTable 12. Estimated effect of target therapies in PedSep-D from logistic regression

eFigure 1. Schematic of study methods

eFigure 2. Heatmap of correlation between clinical variables for phenotyping

eFigure 3. OPTICS plots (N= 404)

eFigure 4. Consensus k clustering results

eFigure 5. Comparison of variables that contribute to clinical phenotypes using Consensus k-means and LCA

eFigure 6. Sensitivity analysis using latent class clustering (N=404), showing probabilities of phenotype assignment.

eFigure 7. Comparison of phenotype membership between Consensus k-means and LCA

eFigure 8. Comparison of select inflammatory cytokines bar graphs across phenotypes

eFigure 9. Alluvial plot showing distribution of phenotypes across baseline OFI (N=404)

eFigure 10. Mortality and organ failure by phenotype over 28 days

eFigure 11. Comparison of day 1 clinical variables that contribute to survival within each phenotype

eFigure 12. Comparison of Cytokine Biomarkers That Contribute to Survival within Each Phenotype

eFigure 13. t-SNE plot of outcome, IPMOF, TAMOF, SMOF, and MAS (N = 404)

eFigure 14. Count of patients (14 therapies)

#### eTable 1. List of all variables collected in parent study

| Variable | Description of variable | Reason for exclusion |
|---|---|---|
| **Demographic** | | |
| Age | | |
| Sex | | |
| Race | | Self-reported race with multiple overlapping groups |
| Ethnicity | | |
| Previous healthy | | |
| Surgery | | |
| **PRISM[a]** | | |
| Low SBP | Lowest Systolic Blood Pressure | |
| High Heart Rate | Highest Heart Rate | |
| Low Temp | Lowest Temperature | |
| High Temp | Highest Temperature | |
| Pupillary Reflex | Number of pupils > 3 mm and fixed | Non-informative[b] |
| GCS | The lowest GCS score | |
| GCS Eye | GCS Eye | High correlation with GCS |
| GCS Verbal | GCS Verbal | High correlation with GCS |
| GCS Motor | GCS Motor | High correlation with GCS |
| Intubate | Intubated at GCS assessment | |
| Low pH | Lowest pH | High missingness |
| Low PaO2 | Lowest PaO2 | High missingness |
| High PCO2 | Highest PCO2 | High missingness |
| Low Total CO2 | Lowest Total CO2 | High missingness |
| High Glucose | Highest Serum Glucose | High missingness |
| High Potassium | Highest Serum Potassium | High missingness |
| High Creatinine | Highest Creatinine | High correlation with Higher Creatinine |
| Low WBC | Lowest WBC | High missingness |
| Low Platelet | Lowest Platelets | |
| High PT | Highest PT | High missingness |
| High PTT | Highest PTT | High missingness |
| High BUN | Highest Blood urea nitrogen | High missingness |
| **Labs** | | |
| High Creatinine | Creatinine | High correlation with Higher Creatinine |

| | | |
|---|---|---|
| Higher Creatinine | Highest value from PRISM High Creatinine and High Creatinine | |
| High PT | PT | High correlation with HigherPT |
| Higher PT | Highest value from PRISM High PT and High PT | High missingness |
| Low Lymphocyte | Absolute lymphocyte count | |
| Low Neutrophil | Absolute neutrophil count | High missingness |
| Low Hemoglobin | Hemoglobin | |
| Low Platelet | Platelet count | |
| High INR | International normalized ratio | High missingness |
| High Bilirubin | Total bilirubin | High missingness |
| High Lipase | Lipase | High missingness |
| High ALT | ALT | High missingness |
| High LDH | LDH | High missingness |
| High Triglycerides | Triglycerides | High missingness |
| **Organ failure** | | |
| OFI Cardio | Cardiovascular failure: Inotrope OR vasopressor infusion requirement. | |
| OFI Pulm | Pulmonary failure: PaO2/FIO2 ratio of < 300 mm Hg AND mechanical ventilator requirement. | |
| OFI Hepatic | Hepatic failure: ALT > 100 U/L AND either bilirubin > 1.0 mg/dL OR INR > 1.5 | |
| OFI Renal | Renal failure: Creatine > 1mg/dL with Oliguria (urine output < 0.5 mL/kg/hr) | |
| OFI Hemat | Hematologic failure: Platelet count < 100,000 cells/µL AND INR > 1.5 | |
| OFI CNS | Central Nervous System failure: Glasgow coma score < 12 in absence of sedatives | |
| OFI | Total number of organ failure | |
| SIRS | Systemic Inflammatory Response Syndrome criteria | |
| **Cytokine** | | |
| CRP | | |
| Ferritin | | |

a. PRISM: Pediatric Risk of Mortality Index. Variables record the worst physiologic values obtained in following 6 hour timeframe: 2 hours prior to ICU admission through 4 hour post ICU admission.
b. All samples had the same values.

#### eTable 2. Missing data (no., %) for demographic and day 1 clinical variables

| Variable | No. of missing data (%) |
|---|---|
| **Demographic** | |
| Age | 0 (0) |
| Sex | 0 (0) |
| Ethnicity | 0 (0) |
| Previous healthy | 0 (0) |
| Surgery | 0 (0) |
| **Organ Dysfunction** | |
| SIRS criteria | 0 (0) |
| OFI | 0 (0) |
| **Inflammation** | |
| C-reactive protein | 4 (0.9) |
| Low Temperature | 2 (0.4) |
| High Temperature | 2 (0.4) |
| ALC | 79 (19.5) |
| Ferritin | 4 (0.9) |
| **Pulmonary** | |
| Pulmonary OFI | 0 (0) |
| Intubation | 30 (7.4) |
| **Cardiovascular or Hemodynamic** | |
| Heart rate | 0 (0) |
| Systolic blood pressure | 1 (0.2) |
| CV OFI | 0 (0) |
| **Renal** | |
| Creatinine | 19 (4.7) |
| Renal OFI | 0 (0) |
| **Hepatic** | |
| Hepatic OFI | 0 (0) |
| **Hematologic** | |
| Hemoglobin | 48 (11.9) |
| Platelets | 36 (8.9) |
| Hematologic OFI | 0 (0) |
| **Other** | |
| Glasgow Coma Scale score | 0 (0) |
| CNS OFI | 0 (0) |

Abbreviations: SIRS, systemic inflammatory response syndrome; OFI, organ failure index; ALC, absolute lymphocyte count; CNS, central nervous system

#### eTable 3. Statistical test results of demographic and day 1 clinical characteristics (n = 404)

| Characteristic[a] | Statistical test p-value | | | | | | |
|---|---|---|---|---|---|---|---|
| | General | Pairwise | | | | | |
| | | PedSep-A vs PedSep-B | PedSep-A vs PedSep-C | PedSep-A vs PedSep-D | PedSep-B vs PedSep-C | PedSep-B vs PedSep-D | PedSep-C vs PedSep-D |
| **Demographic** | | | | | | | |
| Age | <0.001 | <0.001 | <0.001 | <0.001 | 0.023 | 0.917 | 0.206 |
| Sex | 0.014 | 0.017 | 0.934 | 0.392 | 0.364 | 0.964 | 0.964 |
| Ethnicity | 0.027 | 0.400 | 0.400 | 0.130 | 0.400 | 0.570 | 0.180 |
| Previous healthy | <0.001 | <0.001 | <0.001 | <0.001 | 1.000 | 1.000 | 1.000 |
| Surgery | <0.001 | 0.004 | 0.357 | 0.004 | 0.357 | 0.830 | 0.357 |
| **Organ Dysfunction** | | | | | | | |
| SIRS criteria | 0.221 | 0.830 | 1.000 | 1.000 | 0.360 | 1.000 | 0.830 |
| OFI | <0.001 | <0.001 | 0.420 | <0.001 | <0.001 | <0.001 | <0.001 |
| **Inflammation** | | | | | | | |
| C-reactive protein | <0.001 | <0.001 | <0.001 | 0.002 | 0.137 | 0.952 | 0.265 |
| Low Temperature | <0.001 | 0.002 | <0.001 | 0.108 | <0.001 | 0.298 | <0.001 |
| High Temperature | <0.001 | 0.080 | 0.002 | 0.594 | <0.001 | 0.389 | 0.012 |
| ALC | <0.001 | <0.001 | <0.001 | 0.001 | <0.001 | 0.782 | <0.001 |
| Ferritin | <0.001 | <0.001 | <0.001 | <0.001 | 0.001 | <0.001 | 0.187 |
| **Pulmonary** | | | | | | | |
| Pulmonary OFI | <0.001 | 0.319 | <0.001 | 0.2584 | <0.001 | 0.053 | <0.001 |
| Intubation | <0.001 | <0.001 | <0.001 | 1.000 | <0.001 | <0.001 | <0.001 |
| **Cardiovascular or Hemodynamic** | | | | | | | |
| Heart rate | <0.001 | <0.001 | <0.001 | 0.005 | 0.918 | 0.911 | 0.918 |
| Systolic blood pressure | <0.001 | <0.001 | 0.845 | 0.180 | <0.001 | 0.731 | 0.180 |
| CV OFI | <0.001 | <0.001 | <0.001 | <0.001 | 0.057 | 0.151 | 1.000 |
| **Renal** | | | | | | | |
| Creatinine | <0.001 | <0.001 | <0.001 | <0.001 | 0.320 | <0.001 | <0.001 |
| Renal OFI | <0.001 | 1.000 | 1.000 | <0.001 | 1.000 | <0.001 | <0.001 |
| **Hepatic** | | | | | | | |
| Hepatic OFI | <0.001 | 0.066 | 0.034 | <0.001 | 0.818 | 0.006 | 0.007 |
| **Hematologic** | | | | | | | |
| Hemoglobin | 0.001 | 0.043 | 0.797 | 0.014 | 0.044 | 0.719 | 0.014 |
| Platelets | <0.001 | <0.001 | <0.001 | <0.001 | 0.005 | <0.001 | 0.004 |
| Hematologic OFI | <0.001 | 1.000 | 0.004 | <0.001 | 0.014 | <0.001 | <0.001 |
| **Other** | | | | | | | |
| Glasgow Coma Scale score | <0.001 | <0.001 | <0.001 | 0.642 | <0.001 | 0.001 | <0.001 |
| CNS OFI | <0.001 | 0.016 | 0.891 | 0.092 | 0.002 | 0.918 | 0.017 |

Abbreviations: SIRS, systemic inflammatory response syndrome; OFI, organ failure index; ALC, absolute lymphocyte count; CNS, central nervous system

[a] Comparisons across all 4 phenotypes were performed using the Kruskal-Wallis test for continuous variables, the $\chi 2$ test for categorical variables, or the fisher's exact test for cells with less than 5 patients.

#### eTable 4. Statistical output from latent class analysis

| | Statistic[a] | | | | Class size[b] N, (%) | | | | | |
|---|---|---|---|---|---|---|---|---|---|---|
| **Class number** | AIC | BIC | Entropy[a] | Median [IQR] (%) probability of membership | 1 | 2 | 3 | 4 | 5 | 6 |
| **2** | 44875 | 45199 | 0.917 | 100.0[99.9-100.0] | 308(76) | 96(24) | - | - | - | - |
| **3** | 44329 | 44817 | 0.872 | 99.8[97.1-100.0] | 212(52) | 129(32) | 63(16) | - | - | - |
| **4** | 43774 | 44426 | 0.904 | 99.8[97.4-100.0] | 144 (36) | 142(35) | 73(18) | 45(11) | - | - |
| **5** | 38542 | 39359 | 0.999 | 99.9[98.6-100.0] | 146(36) | 114(28) | 87(22) | 34(8) | 23(6) | - |
| **6** | 39540 | 40520 | 0.999 | 99.8[96.9-100.0] | 98(24) | 87(22) | 83(21) | 74(18) | 42(10) | 20(5) |

Abbreviations: AIC, Akaike information criterion; BIC, Bayesian information criteria; IQR, interquartile range

a AIC and BIC are information criteria for comparing models, where lower value suggests a better fit; Entropy is a measure between 0 and 1 measures success of classification, where a value closer to 1 implies a better fit.
b class size shows the number of samples assigned into each cluster, relatively large size of each cluster is preferred.

#### eTable 5. Biomarkers measured at Day 1 by phenotype

| Biomarker[a] | Total | Phenotype | | | |
|---|---|---|---|---|---|
| | | PedSep-A | PedSep-B | PedSep-C | PedSep-D |
| ADAMTS13, % | 71.0 (56.0, 88.0) | 82.5 (65.0, 95.0) | 71.0 (57.0, 89.3) | 69.0 (52.5, 84.5) | 54.0 (38.0, 66.5) |
| SFasLg, pg/ml | 44.9 (29.0, 73.2) | 58.4 (37.2, 84.6) | 43.2 (31.1, 78.1) | 38.0 (25.0, 65.9) | 36.7 (20.9, 49.3) |
| Ex vivo TNF-α, pg/ml | 427.8 (97.0, 1023.3) | 689.7 (347.1, 1049.2) | 331.1 (99.2, 806.8) | 212.3 (35.7, 668.0) | 278.0 (53.0, 1049.2) |
| TNF-α, pg/ml | 74.9 (56.2, 105.7) | 69.0 (51.9, 85.1) | 74.9 (55.4, 101.8) | 76.2 (55.4, 108.6) | 102.2 (81.4, 131.7) |
| sCD163, pg/ml | 294096 (195700, 496348) | 223123 (163123, 323365) | 309800 (185699, 508775) | 345238 (248348, 572766) | 668162 (280407, 897784) |
| IFN-β, pg/ml | 6.4 (6.4, 8.2) | 6.4 (6.4, 7.2) | 6.4 (6.4, 9.9) | 6.4 (6.4, 10.8) | 6.4 (6.4, 6.4) |
| IL-22, pg/ml | 26.0 (20.1, 34.2) | 22.4 (17.8, 29.5) | 28.0 (21.3, 36.9) | 27.1 (20.1, 34.2) | 31.9 (24.8, 49.2) |
| IL-18, pg/ml | 214.0 (113.5, 325.0) | 192.0 (114.2, 306.2) | 227.5 (113.8, 319.5) | 211.5 (93.0, 345.0) | 246 (177.0, 356.0) |
| IL-18BP, pg/ml | 334.0 (174.0, 498.0) | 363.0 (170.2, 562.8) | 315.5 (154.5, 479.2) | 306.0 (189.5, 442.0) | 363 (226.0, 451.5) |
| MIG/CXCL9, pg/ml | 230.0 (144.0, 313.5) | 250.5 (159.0, 323.0) | 238.0 (151.8, 326.8) | 220.5 (121.2, 303.5) | 221.0 (133.5, 284.5) |
| IL-1β, pg/ml | 2.8 (2.4, 3.3) | 2.6 (2.1, 3.2) | 2.8 (2.3, 3.2) | 2.9 (2.4, 3.3) | 2.9 (2.5, 3.3) |
| IL-4, pg/ml | 4.7 (3.5, 6.5) | 4.9 (3.5, 6.3) | 4.9 (3.9, 6.8) | 4.7 (3.5, 6.7) | 4.3 (3.5, 6.4) |
| IL-6, pg/ml | 8.8 (6.5, 19.0) | 6.9 (5.8, 10.0) | 8.7 (6.5, 25.2) | 11.1 (7.0, 27.2) | 16.8 (8.4, 43.8) |
| IL-8, pg/ml | 55.0 (31.4, 108.6) | 41.2 (27.7, 66.7) | 57.1 (34.7, 113.7) | 57.5 (35.1, 127.9) | 123.5 (71.2, 468.6) |
| IL-10, pg/ml | 22.5 (17.5, 33.4) | 19.3 (15.4, 24.6) | 22.5 (18.6, 37.1) | 23.7 (18.1, 37.5) | 32.8 (24.6, 73.3) |
| IL-13, pg/ml | 3.1 (3.1, 3.9) | 3.1 (3.1, 4.2) | 3.1 (3.1, 3.4) | 3.1 (3.1, 4.3) | 3.1 (3.1, 3.4) |
| IL-17A, pg/ml | 12.0 (9.0, 19.0) | 10.0 (7.0, 17.0) | 12.0 (9.0, 19.0) | 16.0 (9.3, 23.0) | 12.0 (9.0, 19.0) |
| IFN-γ, pg/ml | 2.8 (2.8, 2.8) | 2.8 (2.8, 3.0) | 2.8 (2.8, 3.0) | 2.8 (2.8, 3.0) | 2.8 (2.8, 3.2) |
| IP-10/CXCL10, pg/ml | 727.8 (343.9, 1963.7) | 494.0 (287.3, 1725.7) | 705.8 (259.7, 1585.7) | 967.7 (409.0, 2334.1) | 789.5 (466.8, 2109.7) |
| MCP-1/CCL2, pg/ml | 142.5 (70.9, 367.6) | 103.6 (49.7, 197.2) | 169.8 (83.9, 383.3) | 184.4 (88.7, 474.6) | 240.7 (114.5, 1623.0) |
| MIP-1α, pg/ml | 0.6 (0.6, 7.7) | 0.6 (0.6, 0.6) | 0.6 (0.6, 8.1) | 0.6 (0.6, 9.0) | 6.6 (2.8, 16.1) |
| MIP-1β, pg/ml | 45.9 (31.4, 70.4) | 42.4 (28.1, 56.3) | 45.7 (31.9, 73.9) | 44.6 (32.9, 77.8) | 58.4 (47.9, 95.0) |
| MCP-3, pg/ml | 92.4 (92.4, 166.0) | 92.4 (92.4, 147.8) | 92.4 (92.4, 166.0) | 119.5 (92.4, 166.0) | 119.5 (92.4, 180.6) |
| IFN-α2, pg/ml | 125.7 (105.8, 140.2) | 124.3 (105.8, 140.2) | 125.7 (108.8, 144.4) | 125.7 (105.8, 140.2) | 120.0 (105.8, 137.9) |
| IL-1α, pg/ml | 9.4 (9.4, 13.2) | 9.4 (9.4, 9.9) | 9.4 (9.4, 16.4) | 9.4 (9.4, 15.6) | 9.4 (9.4, 13.2) |
| IL-2RA, pg/ml | 378.8 (243.0, 623.2) | 345.2 (237.6, 511.6) | 385.3 (206.6, 683.1) | 380.6 (243.1, 731.5) | 449.7 (307.9, 747.5) |
| IL-3, pg/ml | 612.2 (529.0, 724.4) | 624.4 (496.1, 734.6) | 636.6 (529.0, 724.4) | 612.2 (529.0, 724.4) | 586.4 (496.1, 693.0) |

| | | | | | |
|---|---|---|---|---|---|
| IL-16, pg/ml | 569.8 (410.2, 763.0) | 544.4 (391.2, 677.2) | 590.4 (435.2, 770.2) | 529.4 (382.7, 704.3) | 858.0 (592.6, 1246.4) |
| M-CSF, pg/ml | 30.0 (17.0, 55.3) | 20.7 (14.2, 33.6) | 30.6 (20.3, 54.6) | 34.4 (20.3, 58.9) | 79.9 (46.2, 122.7) |
| SCF, pg/ml | 160.2 (118.8, 244.4) | 141.6 (115.0, 199.9) | 158.2 (113.8, 226.8) | 151.5 (114.2, 238.8) | 326.1 (227.5, 504.1) |
| TRAIL, pg/ml | 36.6 (27.9, 54.2) | 42.9 (32.9, 65.5) | 35.4 (25.4, 54.5) | 35.4 (29.1, 45.4) | 27.9 (24.1, 40.4) |
| CRPH, mg/dL | 9.8 (3.3, 17.1) | 4.3 (1.2, 12.4) | 10.1 (4.8, 19.3) | 14.3 (7.5, 21.7) | 10.7 (3.4, 20.7) |
| Ferritin, ng/mL | 218.0 (98.0, 625.3) | 125.4 (69.8, 207.8) | 223.1 (116.5, 544.2) | 405.5 (176.2, 1485.7) | 610.0 (221.1, 2482.0) |

[a] All biomarkers are measured in the first day of admission. Values in table are summarized as median (IQR)

#### eTable 6. Statistical test of Biomarkers measured at Day 1

| Biomarker[a] | General | Pairwise | | | | | |
|---|---|---|---|---|---|---|---|
| | | PedSep-A vs PedSep-B | PedSep-A vs PedSep-C | PedSep-A vs PedSep-D | PedSep-B vs PedSep-C | PedSep-B vs PedSep-D | PedSep-C vs PedSep-D |
| ADAMTS13 | <0.001 | 0.033 | 0.001 | <0.001 | 1.000 | <0.001 | <0.001 |
| sFasLg | <0.001 | 0.140 | <0.001 | <0.001 | 0.419 | 0.111 | 1.000 |
| Ex vivo TNF-α | <0.001 | 0.002 | <0.001 | 0.061 | 0.506 | 1.000 | 1.000 |
| TNF-α | <0.001 | 0.321 | 0.152 | <0.001 | 1.000 | <0.001 | 0.002 |
| sCD163 | <0.001 | 0.008 | <0.001 | <0.001 | 0.680 | 0.0015 | 0.021 |
| IFN-β | 0.068 | 0.400 | 0.320 | 1.000 | 1.000 | 0.400 | 0.430 |
| IL-22 | <0.001 | 0.005 | 0.004 | <0.001 | 1.000 | 0.270 | 0.139 |
| IL-18 | 0.106 | 1.000 | 1.000 | 0.046 | 1.000 | 1.000 | 0.591 |
| IL-18BP | 0.165 | 0.440 | 0.410 | 1.000 | 1.000 | 1.000 | 1.000 |
| MIG/CXCL9 | 0.144 | 1.000 | 0.250 | 0.520 | 1.000 | 1.000 | 1.000 |
| IL-1β | 0.071 | 1.000 | 0.130 | 0.250 | 1.000 | 1.000 | 1.000 |
| IL-4 | 0.590 | 1.000 | 1.000 | 1.000 | 1.000 | 1.000 | 1.000 |
| IL-6 | <0.001 | 0.020 | <0.001 | <0.001 | 0.759 | 0.023 | 0.258 |
| IL-8 | <0.001 | 0.008 | 0.002 | <0.001 | 1.000 | <0.001 | <0.001 |
| IL-10 | <0.001 | 0.003 | <0.001 | <0.001 | 1.000 | 0.003 | 0.012 |
| IL-13 | 0.824 | 1.000 | 1.000 | 1.000 | 1.000 | 1.000 | 1.000 |
| IL-17A | 0.002 | 0.166 | <0.001 | 0.862 | 0.858 | 1.000 | 0.577 |
| IFN-γ | 0.998 | 1.000 | 1.000 | 1.000 | 1.000 | 1.000 | 1.000 |
| IP-10/CXCL10 | 0.013 | 1.000 | 0.024 | 0.133 | 0.267 | 0.817 | 1.000 |
| MCP-1/CCL2 | <0.001 | 0.003 | <0.001 | <0.001 | 1.000 | 0.079 | 0.371 |
| MIP-1α | <0.001 | <0.001 | <0.001 | <0.001 | 1.000 | 0.006 | 0.007 |
| MIP-1β | <0.001 | 0.168 | 0.093 | <0.001 | 1.000 | 0.037 | 0.084 |
| MCP-3 | 0.309 | 1.000 | 0.930 | 0.660 | 1.000 | 1.000 | 1.000 |
| IFN-α2 | 0.803 | 1.000 | 1.000 | 1.000 | 1.000 | 1.000 | 1.000 |
| IL-1α | 0.500 | 1.000 | 1.000 | 1.000 | 1.000 | 1.000 | 1.000 |
| IL-2RA | 0.021 | 1.000 | 0.462 | 0.007 | 1.000 | 0.565 | 1.000 |
| IL-3 | 0.596 | 1.000 | 1.000 | 1.000 | 1.000 | 1.000 | 1.000 |
| IL-16 | <0.001 | 0.318 | 1.000 | <0.001 | 0.488 | <0.001 | <0.001 |
| M-CSF | <0.001 | <0.001 | <0.001 | <0.001 | 1.000 | <0.001 | <0.001 |
| SCF | <0.001 | 0.760 | 0.850 | <0.001 | 1.000 | <0.001 | <0.001 |
| TRAIL | <0.001 | 0.032 | 0.003 | <0.001 | 1.000 | 0.104 | 0.047 |
| CRPH | <0.001 | <0.001 | <0.001 | 0.002 | 0.137 | 0.952 | 0.265 |
| Ferritin | <0.001 | <0.001 | <0.001 | <0.001 | <0.001 | <0.001 | 0.187 |

[a] All biomarkers are measured in the first day of admission.

#### eTable 7. Statistical Test Results of Subsequent Outcome Characteristics (n = 404)

| Characteristic[a] | Statistical test p-value | | | | | | |
|---|---|---|---|---|---|---|---|
| | General | Pairwise | | | | | |
| | | PedSep-A vs PedSep-B | PedSep-A vs PedSep-C | PedSep-A vs PedSep-D | PedSep-B vs PedSep-C | PedSep-B vs PedSep-D | PedSep-C vs PedSep-D |
| **MOF Empirical Phenotypes** | | | | | | | |
| SMOF | <0.001 | 1.000 | 1.000 | 0.003 | 1.000 | 0.008 | 0.027 |
| TAMOF | <0.001 | 0.017 | 0.173 | <0.001 | 0.321 | <0.001 | <0.001 |
| IPMOF | <0.001 | 0.00083 | 0.064 | <0.001 | 0.446 | 0.446 | 0.064 |
| MAS | <0.001 | 0.240 | 0.390 | <0.001 | 0.680 | <0.001 | <0.001 |
| NPMOF | <0.001 | 1.000 | 0.489 | <0.001 | 1.000 | <0.001 | 0.003 |
| **Infections** | | | | | | | |
| Bacterial infection | 0.440 | 1.000 | 1.000 | 1.000 | 1.000 | 1.000 | 1.000 |
| Viral infection | <0.001 | 0.002 | 0.002 | 0.002 | 1.000 | 1.000 | 1.000 |
| Fungal infection | 0.003 | 1.000 | 1.000 | 0.140 | 1.000 | 0.510 | 0.190 |
| Culture negative | 0.049 | 0.096 | 0.432 | 0.336 | 1.000 | 1.000 | 1.000 |
| **Sites** | | | | | | | |
| Blood | <0.001 | 1.000 | 0.023 | 0.023 | 0.023 | 0.019 | 1.000 |
| Lung | 0.006 | 0.634 | 0.245 | 0.634 | 0.014 | 0.185 | 0.963 |
| Urine | 0.644 | 1.000 | 1.000 | 1.000 | 1.000 | 1.000 | 1.000 |
| **Organ Support** | | | | | | | |
| MechVent | <0.001 | 1.000 | <0.001 | 0.302 | <0.001 | 0.302 | 0.013 |
| ECMO | 0.006 | 0.629 | 0.983 | 0.015 | 0.983 | 0.629 | 0.112 |
| CRRT | <0.001 | 0.067 | 0.067 | <0.001 | 1.000 | <0.001 | <0.001 |
| **Anti-inflammatory therapies** | | | | | | | |
| Decadron | <0.001 | 0.057 | <0.001 | 0.002 | 0.561 | 0.561 | 0.926 |
| Methylprednisone | 0.009 | 0.034 | 0.086 | 0.203 | 1.000 | 1.000 | 1.000 |
| IVIG | <0.001 | 0.300 | 0.002 | <0.001 | 0.231 | 0.054 | 0.496 |
| Plasma exchange | <0.001 | 1.000 | 1.000 | 0.002 | 1.000 | 0.006 | 0.002 |
| **Outcome** | | | | | | | |
| Length of Stay | <0.001 | 0.365 | 0.010 | 0.052 | 0.003 | 0.259 | <0.001 |
| Mortality | <0.001 | 0.015 | 0.023 | <0.001 | 0.826 | 0.006 | 0.002 |

Abbreviations: SMOF, sequential liver failure associated multiple organ failure; TAMOF, thrombocytopenia associated multiple organ failure; IPMOF, immunoparalysis associated multiple organ failure; MAS, macrophage activation syndrome; NPMOF, new or progressive multiple organ failure; IQR, interquartile range; MechVent, Mechanical Ventilation; ECMO, Extracorporeal Membrane Oxygenation; CRRT, Continuous Renal Replacement Therapies; IVIG, intravenous gamma globulin

[a] Comparisons across all 4 phenotypes were performed using the Kruskal-Wallis test for continuous variables, the χ2 test for categorical variables, or the fisher's exact test for cells with less than 5 patient

#### eTable 8. Statistical test result of association between day 1 characteristics and mortality

| | Phenotype[c] | | | |
|---|---|---|---|---|
| Characteristic | PedSep-A | PedSep-B | PedSep-C | PedSep-D |
| **Demographic** | | | | |
| Age | 0.45 | 0.39 | 0.55 | 0.78 |
| Sex | 1.00 | 1.00 | 0.80 | 0.26 |
| Ethnicity | 0.58 | 0.67 | 0.55 | 0.61 |
| Previous healthy | 1.00 | 0.58 | 0.89 | 0.02[b] |
| Surgery | 1.00 | 0.56 | 1.00 | 0.77 |
| **Organ Dysfunction** | | | | |
| SIRS criteria | 0.86 | 0.85 | 0.74 | 0.20 |
| OFI | 0.75 | 0.67 | 0.23 | 0.20 |
| **Inflammation** | | | | |
| C-reactive protein | 0.34 | 0.13 | 0.52 | 0.68 |
| Low Temperature | 0.45 | 0.44 | 0.61 | 0.30 |
| High Temperature | 0.71 | 0.4 | 0.02[b] | 0.80 |
| ALC | 0.89 | 0.35 | 0.02[b] | 0.23 |
| Ferritin | 0.64 | 0.95 | 0.03[a] | <0.01[a] |
| **Pulmonary** | | | | |
| Pulmonary OFI | 1.00 | 1.00 | 0.06 | 0.33 |
| Intubation | 0.92 | 0.61 | 1.00 | 0.86 |
| **Cardiovascular or Hemodynamic** | | | | |
| Heart rate | 0.57 | 0.53 | 0.07 | 0.95 |
| Systolic blood pressure | 0.96 | 1.00 | <0.01[a] | 0.08 |
| Cardiovascular OFI | 1.00 | 0.74 | 0.45 | 0.08 |
| **Renal** | | | | |
| Creatinine | 0.08 | 0.14 | 0.44 | 0.86 |
| Renal OFI | - | - | - | 0.34 |
| **Hepatic** | | | | |
| Hepatic OFI | 1.00 | 0.54 | 0.67 | 0.87 |
| **Hematologic** | | | | |
| Hemoglobin | 0.55 | 0.33 | 0.47 | 0.91 |
| Platelets | 0.34 | 0.89 | 0.16 | 0.26 |
| Hematologic OFI | - | - | 1.00 | 0.58 |
| **Other** | | | | |
| Glasgow Coma Scale score | 0.32 | 0.89 | 0.39 | 0.30 |
| CNS OFI | 0.01[a] | 0.62 | 0.89 | 1.00 |

Abbreviations: SIRS, systemic inflammatory response syndrome; OFI, organ failure index; ALC, absolute lymphocyte count; CNS, central nervous system

[a] Non-survivors have significantly higher value (proportion) of characteristic than survivors.

[b] Non-survivors have significantly lower value (proportion) of characteristic than survivors.

[c] Kruskal-Wallis or chi-square p-value, as appropriate, comparing non-survivors and survivors.

Cells without p-value result from 0 counts of individuals in tested group.

#### eTable 9. Statistical test result of association between biomarkers and mortality

| Biomarker | Phenotype[c] | | | |
|---|---|---|---|---|
| | PedSep-A | PedSep-B | PedSep-C | PedSep-D |
| CRPH | 0.34 | 0.13 | 0.52 | 0.68 |
| Ferritin | 0.64 | 0.95 | 0.03[a] | <0.01[a] |
| ADAMTS13 | 0.25 | 0.43 | 0.13 | 0.84 |
| sFasLg | 0.35 | 0.36 | 0.76 | 0.04[b] |
| Ex vivo TNF-α | 0.31 | 0.71 | 0.88 | 1.00 |
| TNF-α | 0.04[b] | 0.68 | 0.80 | 0.74 |
| sCD163 | 0.87 | 0.52 | 0.08 | 0.14 |
| IFN-β | 0.32 | 0.17 | 0.54 | 0.34 |
| IL-22 | 0.96 | 0.61 | 0.76 | 0.58 |
| IL-18 | 0.06 | 0.78 | 0.51 | 0.03[b] |
| IL-18BP | 0.37 | 0.38 | 0.64 | 0.06 |
| MIG/CXCL9 | 0.06 | 0.06 | 0.09 | 0.23 |
| IL-1β | 0.51 | 0.30 | 0.61 | 0.82 |
| IL-4 | 0.71 | 0.10 | 0.75 | 0.96 |
| IL-6 | 0.16 | 0.04[a] | 0.26 | 0.99 |
| IL-8 | 0.16 | 0.02[a] | <0.01[a] | 0.01[a] |
| IL-10 | 0.33 | 0.75 | 0.11 | <0.01[a] |
| IL-13 | 0.88 | 0.40 | 0.58 | 0.66 |
| IL-17A | 0.12 | 0.75 | 0.44 | 0.71 |
| IFN-γ | 0.25 | 0.06 | 0.38 | 0.15 |
| IP-10/CXCL10 | 0.29 | 0.08 | 0.07 | 0.21 |
| MCP-1/CCL2 | 0.36 | <0.01[a] | 0.06 | 0.11 |
| MIP-1α | 0.36 | 0.07 | 0.14 | <0.01[a] |
| MIP-1β | 0.12 | 0.06 | 0.38 | 0.10 |
| MCP-3 | 0.03[a] | 0.90 | 0.07 | 0.10 |
| IFN-α2 | 0.19 | 0.81 | 0.65 | 0.46 |
| IL-1α | 0.18 | 0.20 | 0.54 | 0.68 |
| IL-2RA | <0.01[b] | 0.64 | 0.81 | 0.71 |
| IL-3 | 0.73 | 0.19 | 0.82 | 0.92 |
| IL-16 | 0.56 | 0.63 | 0.12 | 0.44 |
| M-CSF | 0.22 | 0.17 | 0.18 | 0.61 |
| SCF | 0.17 | 0.73 | 0.16 | 0.10 |
| TRAIL | 0.72 | 0.73 | 0.98 | 0.61 |

[a] Non-survivors have significantly higher value of biomarker than survivors.
[b] Non-survivors have significantly lower value of biomarker than survivors.
[c] Kruskal-Wallis p-value comparing non-survivors and survivors.

#### eTable 10. Statistical test result of 44 candidate therapies

| Therapy | General | PedSep-A | PedSep-B | PedSep-C | PedSep-D |
|---|---|---|---|---|---|
| ANAKINRA, p-value (No.) | 0.017 (5) | - (0) | 1 (1) | 0.055 (3) | 0.367 (1) |
| BECLOMETHASONE, p-value (No.) | 1 (2) | - (0) | - (0) | 1 (2) | - (0) |
| BORTEZOMIB, p-value (No.) | 1 (1) | - (0) | - (0) | - (0) | 1 (1) |
| CAMPATH, p-value (No.) | 1 (2) | - (0) | 1 (1) | 1 (1) | - (0) |
| CARBOPLATIN, p-value (No.) | 1 (1) | 1 (1) | - (0) | - (0) | - (0) |
| CELLCEPT, p-value (No.) | 1 (3) | - (0) | - (0) | 1 (1) | 0.526 (2) |
| CISPLATIN, p-value (No.) | 1 (1) | - (0) | - (0) | 1 (1) | - (0) |
| CYCLOPHOSPHAMIDE, p-value (No.) | 1 (1) | - (0) | - (0) | 1 (1) | - (0) |
| CYCLOSPORINE, p-value (No.) | 0.017 (2) | - (0) | - (0) | 0.147 (1) | 0.367 (1) |
| CYTARABINE, p-value (No.) | 0.342 (3) | - (0) | - (0) | 0.274 (2) | 1 (1) |
| CYTOGAM, p-value (No.) | 0.130 (1) | - (0) | - (0) | - (0) | 0.367 (1) |
| DAUNORUBICIN, p-value (No.) | 1 (1) | - (0) | - (0) | - (0) | 1 (1) |
| DECADRON, p-value (No.) | 0.580 (94) | 1 (50) | 1 (22) | 0.004 (14) | 1 (8) |
| DOXORUBICIN, p-value (No.) | 1 (1) | - (0) | - (0) | 1 (1) | - (0) |
| ENBREL, p-value (No.) | 0.130 (1) | - (0) | - (0) | 0.147 (1) | - (0) |
| EPOETIN ALFA, p-value (No.) | 1 (1) | - (0) | - (0) | - (0) | 1 (1) |
| ETANERCEPT, p-value (No.) | 1 (1) | 1 (1) | - (0) | - (0) | - (0) |
| ETOPOSIDE, p-value (No.) | 0.128 (5) | 1 (1) | - (0) | 0.020 (2) | 0.526 (2) |
| FLUDROCORTISONE, p-value (No.) | 1 (1) | 1 (1) | - (0) | - (0) | - (0) |
| FLUOROURACIL, p-value (No.) | 1 (1) | - (0) | - (0) | 1 (1) | - (0) |
| HYDROCORTISONE, p-value (No.) | 0.061 (172) | 1 (36) | 1 (48) | 1 (50) | 0.724 (38) |
| HYDROXYCHLOROQUINE, p-value (No.) | 1 (2) | 1 (1) | - (0) | 1 (1) | - (0) |
| HYDROXYUREA, p-value (No.) | 0.017 (2) | - (0) | - (0) | - (0) | 0.130 (2) |
| IMMUNOGLOBULIN G, p-value (No.) | 0.001 (51) | 1 (6) | 0.113 (10) | 0.025 (19) | 0.537 (16) |
| INFLIXIMAB, p-value (No.) | 0.130 (1) | - (0) | - (0) | 0.147 (1) | - (0) |
| METHYLPREDNISOLONE, p-value (No.) | 0.862 (117) | 1 (54) | 0.714 (23) | 0.004 (24) | 0.754 (16) |
| MYCOPHENOLATE, p-value (No.) | 0.001 (8) | - (0) | 1 (1) | 0.009 (4) | 0.546 (3) |
| NEUPOGEN, p-value (No.) | 0.001 (23) | - (0) | 0.241 (2) | 0.012 (12) | 0.708 (9) |
| PREDNISOLONE, p-value (No.) | 0.781 (32) | 1 (14) | 1 (5) | 0.612 (9) | 1 (4) |
| PROGRAF, p-value (No.) | 0.243 (2) | - (0) | - (0) | 0.274 (2) | - (0) |
| PULMICORT, p-value (No.) | 1 (3) | 1 (1) | 1 (1) | 1 (1) | - (0) |
| RASBURICASE, p-value (No.) | 1 (1) | - (0) | - (0) | - (0) | 1 (1) |
| RITUXIMAB, p-value (No.) | 1 (2) | 1 (1) | 1 (1) | - (0) | - (0) |
| SARGRAMOSTIM, p-value (No.) | 0.130 (1) | - (0) | - (0) | - (0) | 0.367 (1) |
| SIROLIMUS, p-value (No.) | 1 (2) | - (0) | - (0) | 1 (1) | 1 (1) |
| SYMBICORT, p-value (No.) | 1 (1) | 1 (1) | - (0) | - (0) | - (0) |
| TACROLIMUS, p-value (No.) | 0.042 (16) | 1 (1) | 1 (3) | 0.021 (5) | 1 (7) |
| THYMOGLOBULIN, p-value (No.) | 0.130 (1) | - (0) | - (0) | 0.147 (1) | - (0) |
| TOCILIZUMAB, p-value (No.) | 1 (1) | - (0) | - (0) | 1 (1) | - (0) |
| VINCRISTINE, p-value (No.) | 1 (1) | - (0) | - (0) | 1 (1) | - (0) |
| Plasma exchange, p-value (No.) | 0.047 (25) | 1 (5) | 0.067 (4) | 1 (4) | 1 (12) |
| MechVent, p-value (No.) | 0.10 (366) | 1 (134) | 1 (101) | 0.032 (79) | 1 (52) |
| ECMO, p-value (No.) | 0.001 (30) | 0.11 (5) | 0.001 (9) | 0.110 (6) | 0.073 (10) |
| CRRT, p-value (No.) | 0.001 (52) | 1 (1) | 0.003 (7) | 0.14 (7) | 0.23 (37) |

Values of 2-6 columns of table present p values from statistical tests and number of patients treated by each therapy in each phenotype. A p value less than 0.05 indicates a significant association between individual therapy and survival. “-” indicates no patient from a specific phenotype treated by this therapy. Therapies are selected for combination effect analysis if a significant association is detected in patients from at least one of four phenotypes or general population.

#### eTable 11 Common Anti-inflammatory/Immune Medications Duration median [IQR]

| | Frequency of Use on Study n/N (%) | Duration |
|---|---|---|
| Any Immune Medication | 306/404 (76%) | -- |
| Methylprednisolone | 117/306 (38%) | 5 [2, 7] |
| Dexamethasone | 94/306 (30%) | 1 [1, 3] |
| Immunoglobulin G | 51/306 (17%) | 1 [0, 2] |
| G-CSF (Granulocyte colony stimulating factor) | 23/306 (8%) | 7 [4, 15] |
| Tacrolimus | 18/306 (6%) | 9.5 [3, 18] |
| Mycophenolate | 8/306 (3%) | 4 [2, 7] |
| Anakinra | 5/306 (2%) | 11 [10, 16] |
| Etoposide | 5/306 (2%) | 6 [3, 17] |

Medication administrations were recorded on study Days 0-28 inclusive. Medication start and stop dates were truncated Day 0 and Day 28 or discharge, whichever came first, respectively. Duration is defined as the sum of calendar days on study the patient was receiving immune medications. Each medication duration summary only includes patients receiving the medication. Median [Q1, Q3] are reported.

#### eTable 12. Estimated effect of target therapies in PedSep-D from logistic regression

| | Odds ratio | Lower bound | Upper bound | p-value |
|---|---|---|---|---|
| Univariate model | | | | |
| DECADRON | 0.96 | 0.2 | 4.6 | 0.96 |
| METHYLPREDNISONE | 1.43 | 0.4 | 5.08 | 0.58 |
| union of DECADRON and METHYLPREDNISONE | 1.88 | 0.56 | 6.23 | 0.31 |
| IVIG | 0.64 | 0.19 | 2.23 | 0.48 |
| ECMO | 0.3 | 0.07 | 10.12 | 0.09 |
| Plasma Exchange | 1.02 | 0.25 | 4.12 | 0.98 |
| Multivariate model | | | | |
| DECADRON | 0.42 | 0.06 | 2.7 | 0.35 |
| IVIG | 0.42 | 0.11 | 1.6 | 0.2 |
| DECADRON×IVIG | $3.72\times10^{7}$ | $6.32\times10^{-7}$ | inf | 0.99 |
| Multivariate model | | | | |
| METHYLPREDNISOLONE | 0.51 | 0.1 | 2.61 | 0.42 |
| IVIG | 0.21 | 0.03 | 1 | 0.06 |
| METHYLPREDNISOLONE ×IVIG | 23.31 | 1.43 | $7.60\times10^{2}$ | 0.04 |
| Multivariate model | | | | |
| Union of DECADRON and METHYLPREDNISOLONE | 0.69 | 0.15 | 3.06 | 0.62 |
| IVIG | 0.14 | 0.02 | 0.82 | 0.04 |
| Union of DECADRON and METHYLPREDNISOLONE ×IVIG | 30.6 | 1.88 | $1.05\times10^{3}$ | 0.03 |
| Multivariate model | | | | |
| ECMO | 0.16 | 0.02 | 0.91 | 0.05 |
| Plasma Exchange | 0.68 | 0.14 | 3.88 | 0.64 |
| ECMO × Plasma Exchange | 7.33 | 0.28 | $3.06\times10^{2}$ | 0.24 |

Target therapies are selected based on results from Elastic net analysis of PedSep-D phenotype. Univariate model fit a model with each therapy as predictor and survival status as outcome. Multivariate model fit a model with two therapies of interest and their interaction term as predictors and survival status as outcome.

**eFigure 1. Schematic of study**

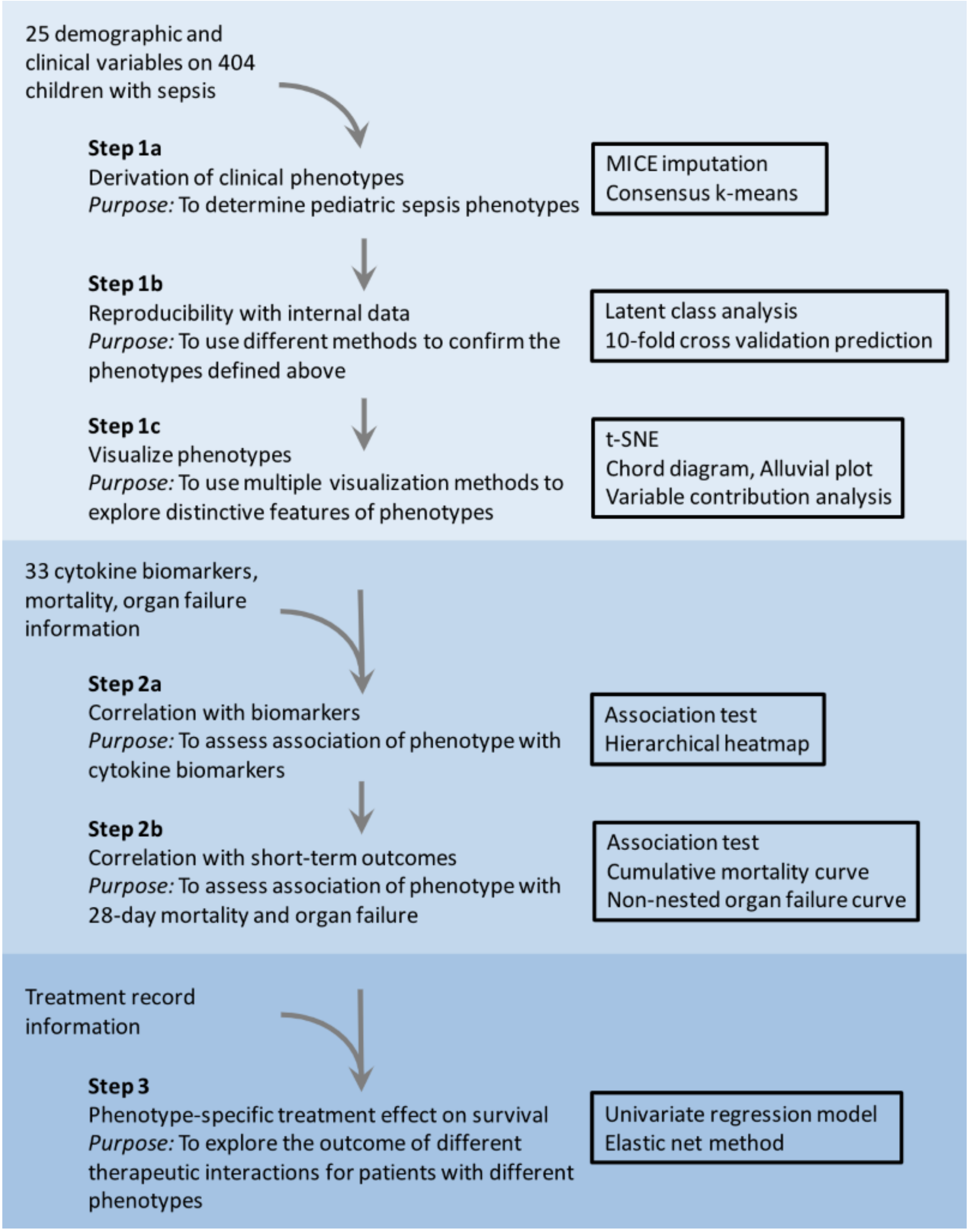

**eFigure 2. Heatmap of correlation between clinical variables for phenotyping**

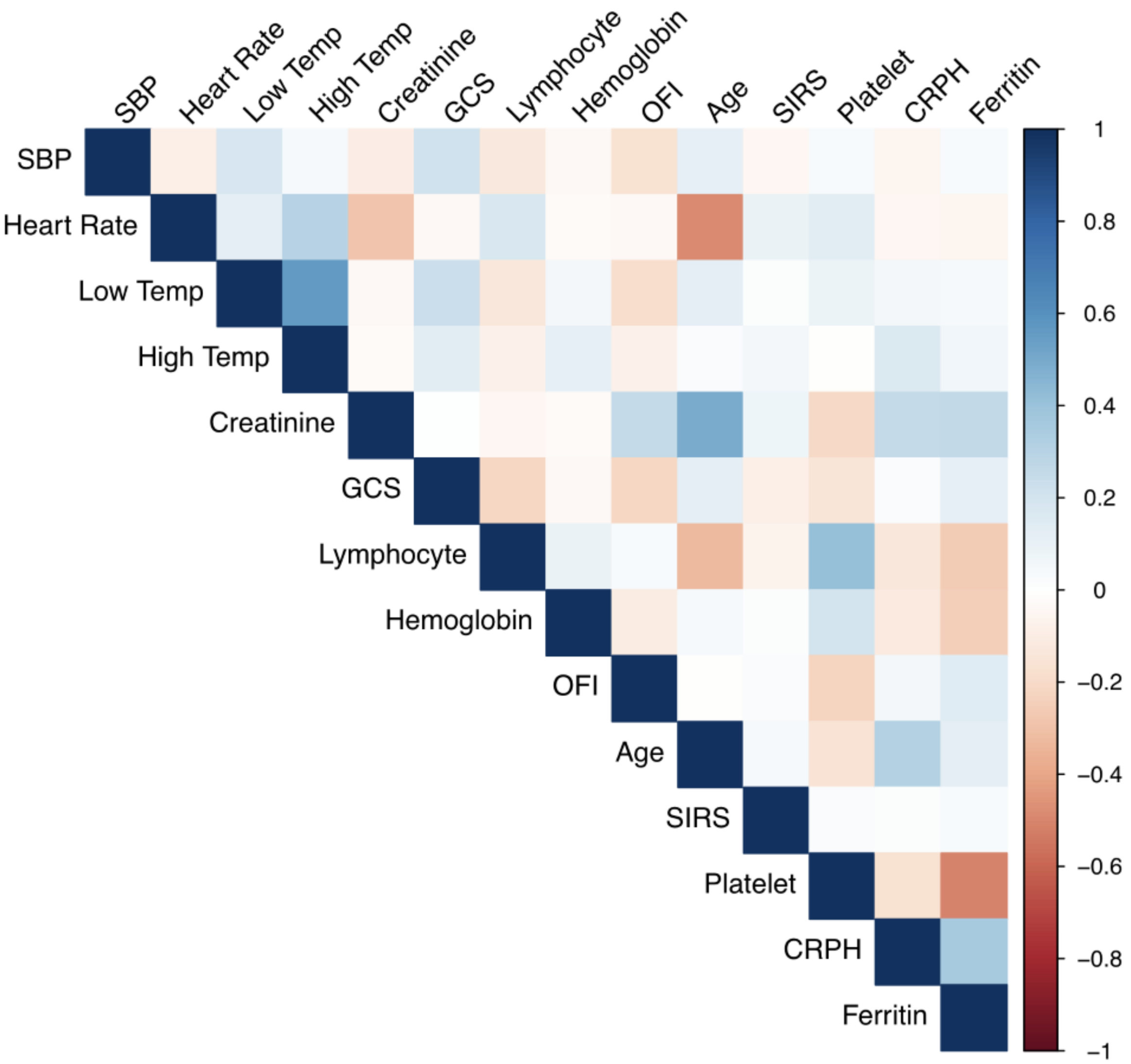


Abbreviations: SBP: Systolic blood pressure; Temp: temperature; GCS: Glasgow coma scale score; OFI, organ failure index; SIRS, systemic inflammatory response syndrome; CRPH, C-reactive protein

Heatmap shows darker color (red or blue) when the Spearman rank order correlation coefficient is greater in positive or negative direction.

#### eFigure 3. OPTICS plot (N= 404)

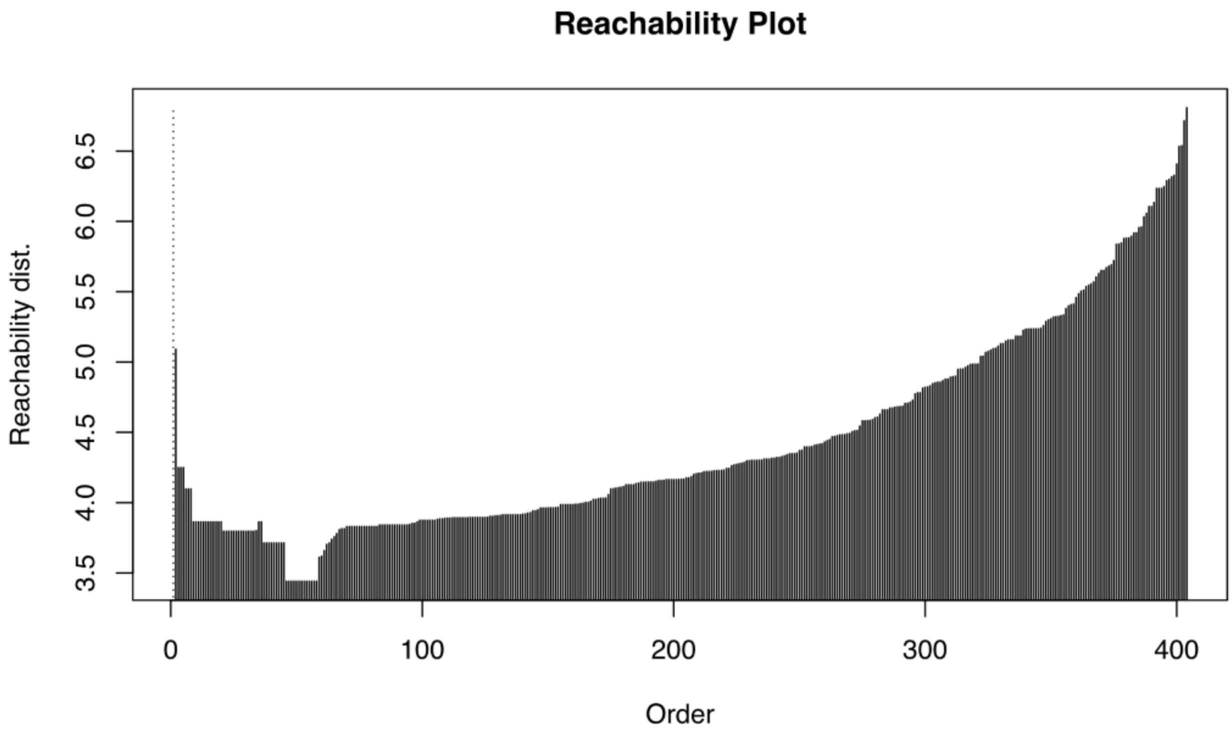


Interpretive example: The OPTICS plot is a figure with the ordering of the patients on the x-axis and the reachability distance on the y-axis. The reachability-distance of two points (samples) is either the distance between them, or the core distance of the core point, whichever is bigger. In our case, the plot shows a smooth rise in reachability distance (as opposed to well demarcated sets). This implies that a partitioning approach such as consensus K means clustering is the preferred statistical algorithm, as opposed to a clustering approach such as hierarchical clustering.

### eFigure 4. Consensus k clustering results

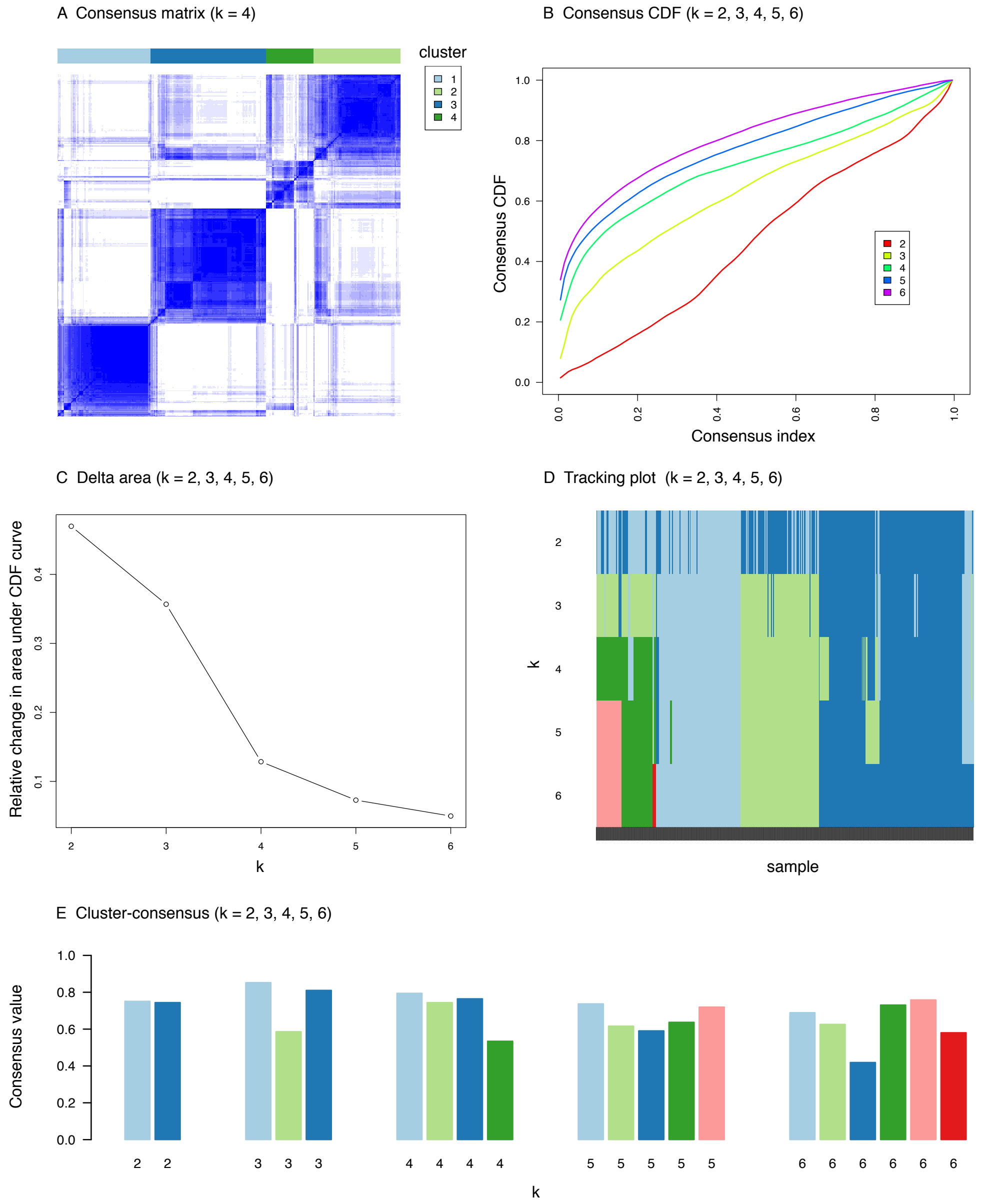


(A) The consensus matrices have patients as both rows and columns. Each consensus value is the frequency the two patients are assigned to the same phenotype among 1000 iterations. It ranges from 0 (white) to 1 (dark blue). This heat map shows a good partition of patients when k=4, where a clear separation between blue and white chunks is observed. (B) Consensus CDF plot shows the cumulative distribution functions of the consensus matrix for each k (indicated by colors), estimated by a histogram of 100 bins. This figure is used to determine at what number of clusters (i.e. k) the CDF reaches an approximate maximum; thus, consensus and cluster confidence is at a maximum at this k. It is usually used together with the Delta area plot to determine the optimal k. (C) Delta area plot shows the relative change in area under the CDF curve comparing k and k − 1. For k = 2, there is no k -1, so the total area under the curve rather than the relative increase is plotted. This plot allows one to determine the relative increase in consensus and determine k at which there is no appreciable increase. Usually, an “elbow” is one of the indicators of the optimal k. In our case, the elbow happens when k = 4, where increasing k makes little contribution. (D) Tracking plot shows the cluster assignment of patients (columns) for each k (rows) by color. The colors correspond to the colors of the consensus matrix class assignments. Each column indicates patient. This plot indicates patient cluster membership change if one were to use k=2, k=3, k=4, k=5, or k=6. Clusters with an abundance of unstable members (changing colors within a column) suggest an unstable cluster. (E) Cluster-consensus plot shows the cluster-consensus value of clusters at k=2, k=3, k=4, k=5, or k = 6. This is the mean of all pairwise consensus values between a cluster’s members. Cluster is indicated by color following the same color scheme as the cluster matrices and tracking plots. The bars are grouped by k which is marked on the horizontal axis. High values indicate a cluster has high stability and low values indicate a cluster has low stability. We used 0.5 as a cut off for diagnostic purposes, where consensus values of all four clusters are higher than this threshold.

#### eFigure 5. Comparison of Variables between consensus k means clustering and LCA

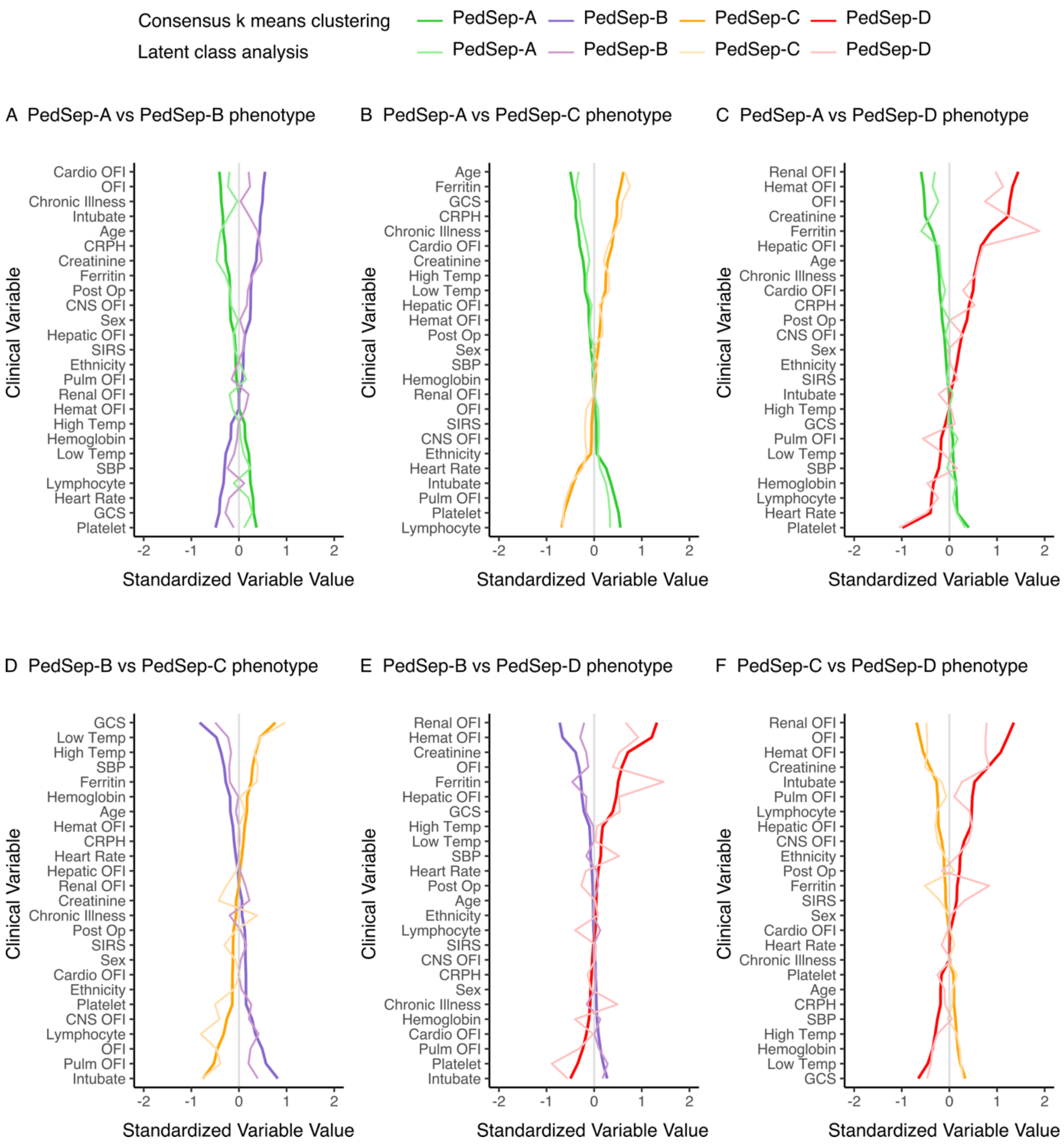


–

Comparisons of variables that contribute to 24-hour clinical phenotypes using consensus k means clustering and latent class analysis (LCA). In all panels, the variables are standardized such that all means are scaled to 0 and SDs to 1. A value of 1 for the standardized variable value (x-axis) signifies that the mean value for the phenotype was 1 SD higher than the mean value for both phenotypes shown in the graph as a whole. CNS - central nervous system; CRP - C-reactive protein; GCS - Glasgow Coma Scale; Hemat - Hematologic; Intubate- Intubation with endotracheal tube; OFI- organ failure index; Post Op - post-surgery; Pulm- pulmonary; Temp- temperature; SBP- systolic blood pressure; Chronic illness – not previously healthy; Ethnicity – higher number with more non-Hispanic; Sex – higher with more males in group.

#### eFigure 6. Sensitivity analysis using latent class clustering (N=404), showing probabilities of PedSep-A, B, C, and D phenotype assignment.

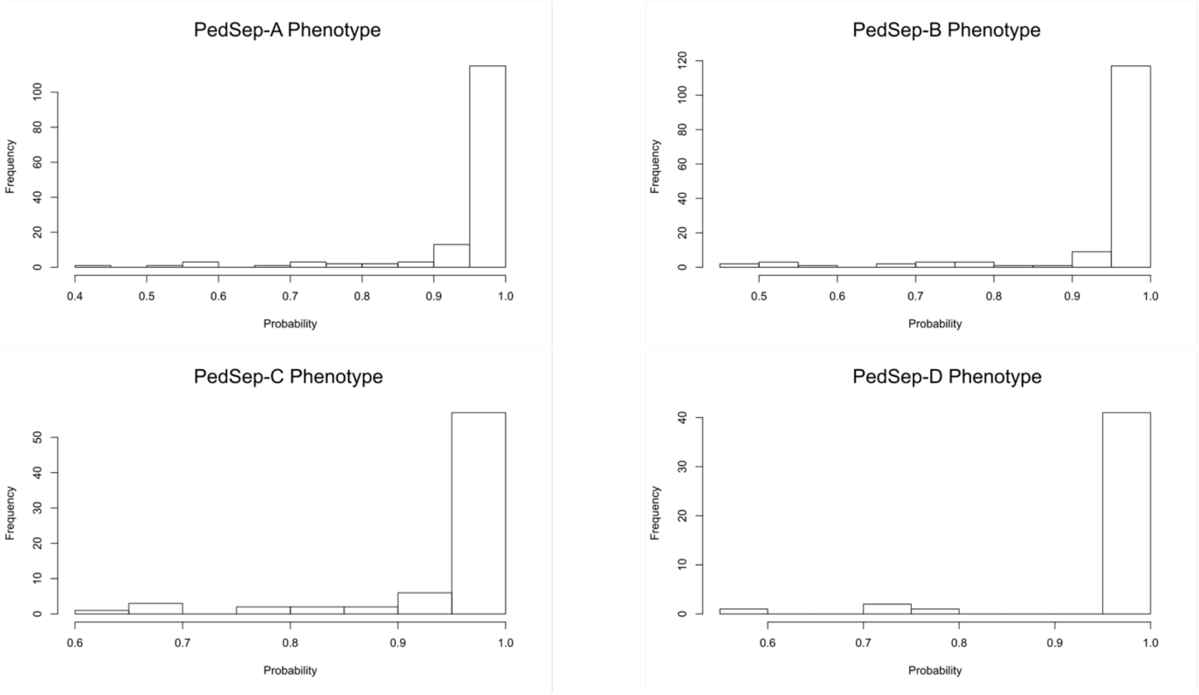


*Interpretive example*: Using latent class analysis to derive phenotypes (called clusters in this output), histograms of within phenotype probability demonstrated that members have high probability of being a phenotype member (>0.9).

#### eFigure 7. Comparison of phenotype membership between Consensus k-means and LCA

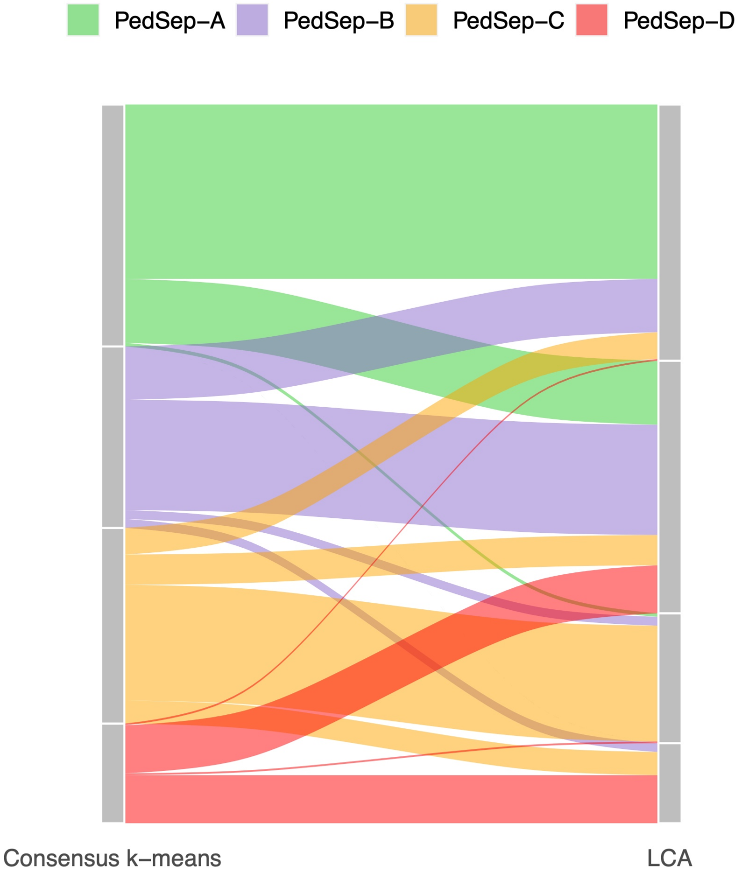


This alluvial diagram shows the phenotype membership difference of 404 patients between Consensus k-means and LCA. The blocks on the two sides represent four phenotypes identified by Consensus k-means and LCA, separately. The streams are colored by phenotypes defined by Consensus k-means. The stream fields between two blocks represent changes in the patient phenotypes assignment in two methods. The height of a block represents the size of the phenotype and the height of a stream field represents the size of the patients contained in both phenotypes connected by the stream field. We observed a consistency of phenotype membership between two cluster methods. While a subset of patients is reassigned in the confirmatory LCA method, the majority of assignment remain robust to method of clustering.

#### eFigure 8. Inflammatory Cytokines Across PedSep A, B, C, and D

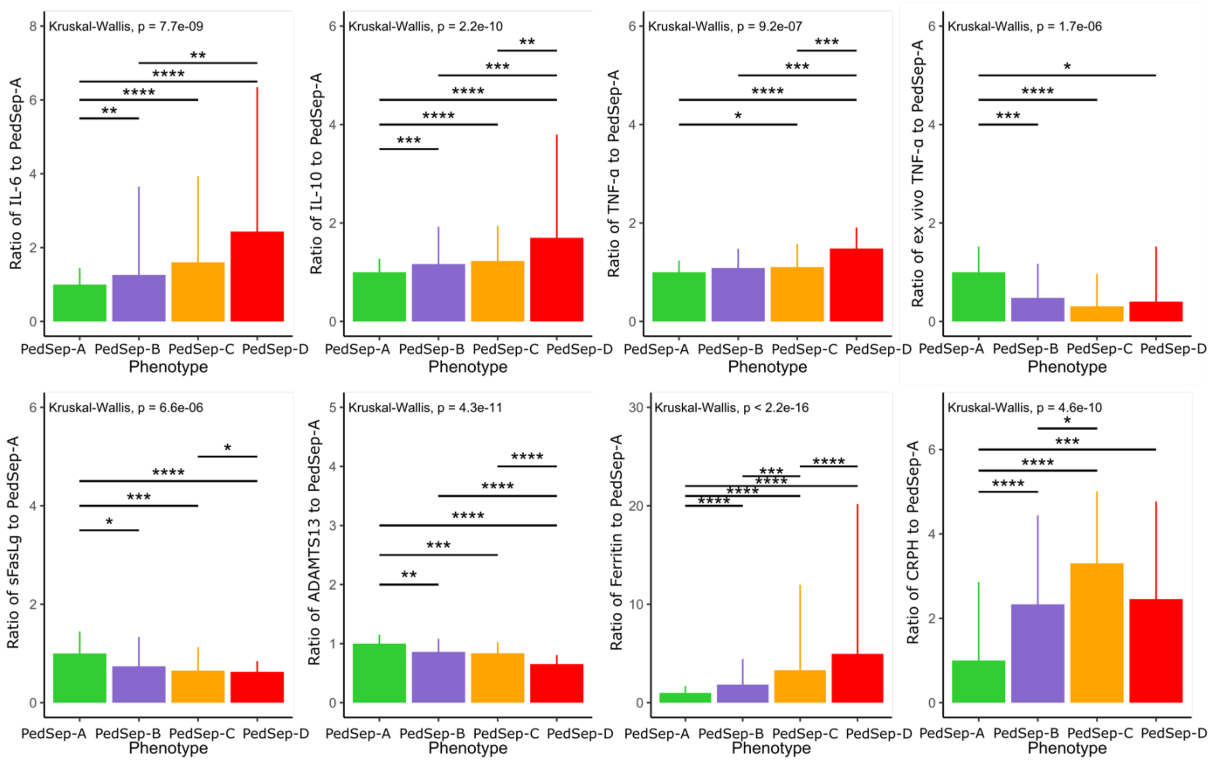


Ratio of each biomarker was calculated as the cytokine value standardized by the median value for the Ped-α phenotype (reference group). All comparisons within data sets across phenotypes were significant (*P* < .001). Errors bars indicate the upper bound of the interquartile range of the biomarker standardized by the median value for the PedSep-A phenotype. Inflammatory cytokines IL-6, IL-10, and TNF measured at baseline were greater in the Ped-C phenotype (orange) and Ped-D phenotype (red) compared with the Ped-A phenotype (green), suggesting a predominantly hyperinflammatory response. TNF indicates tumor necrosis factor.

#### eFigure 9. Alluvial plot showing distribution of PedSep-A,B,C,D and across baseline OFI (N=404)

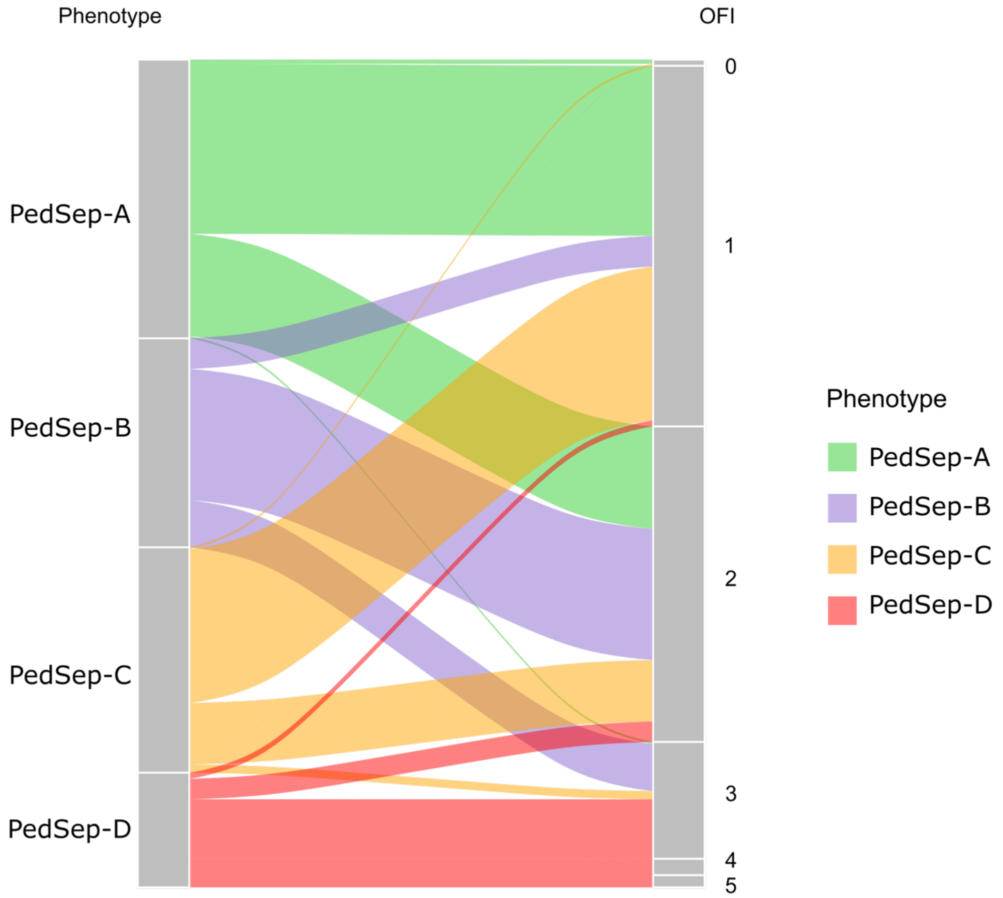


In these alluvial plots, phenotype members are shown by color on the left column and distribute across OFI in the right column. In general, most of PedSep-A and PedSep-C phenotype distributed across lower OFI (OFI less or equal to 2), while phenotype PedSep-B and PedSep-D distributed across higher OFI (OFI is higher or equal to 2 for PedSep-B, OFI is higher or equal to 3 for PedSep-D).

#### eFigure 10. Short-term Mortality and Organ Failure by Phenotype

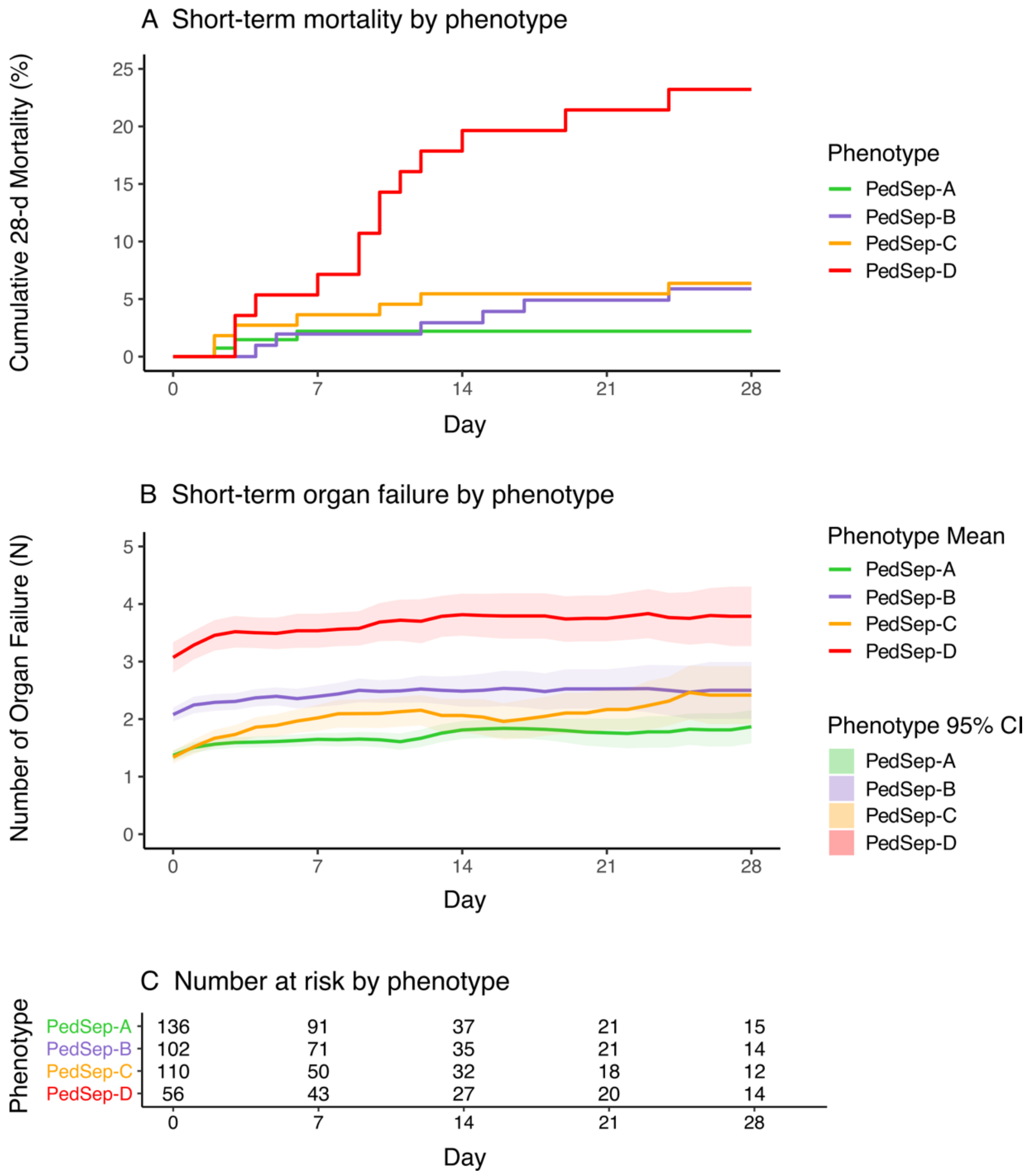


Both short-term mortality (panel A) and organ failure (panel B) show significant differences by phenotype (P < .001). The mean numbers of organ failures and 95% confidence intervals (CI) are calculated each day by non-nested observation. Panel C shows number of people in the ICU at day 0, 7, 14, 21, and 28.

#### eFigure 11. Comparison of Day 1 Variables That Contribute to Survival in PedSep-A, B, C, and D

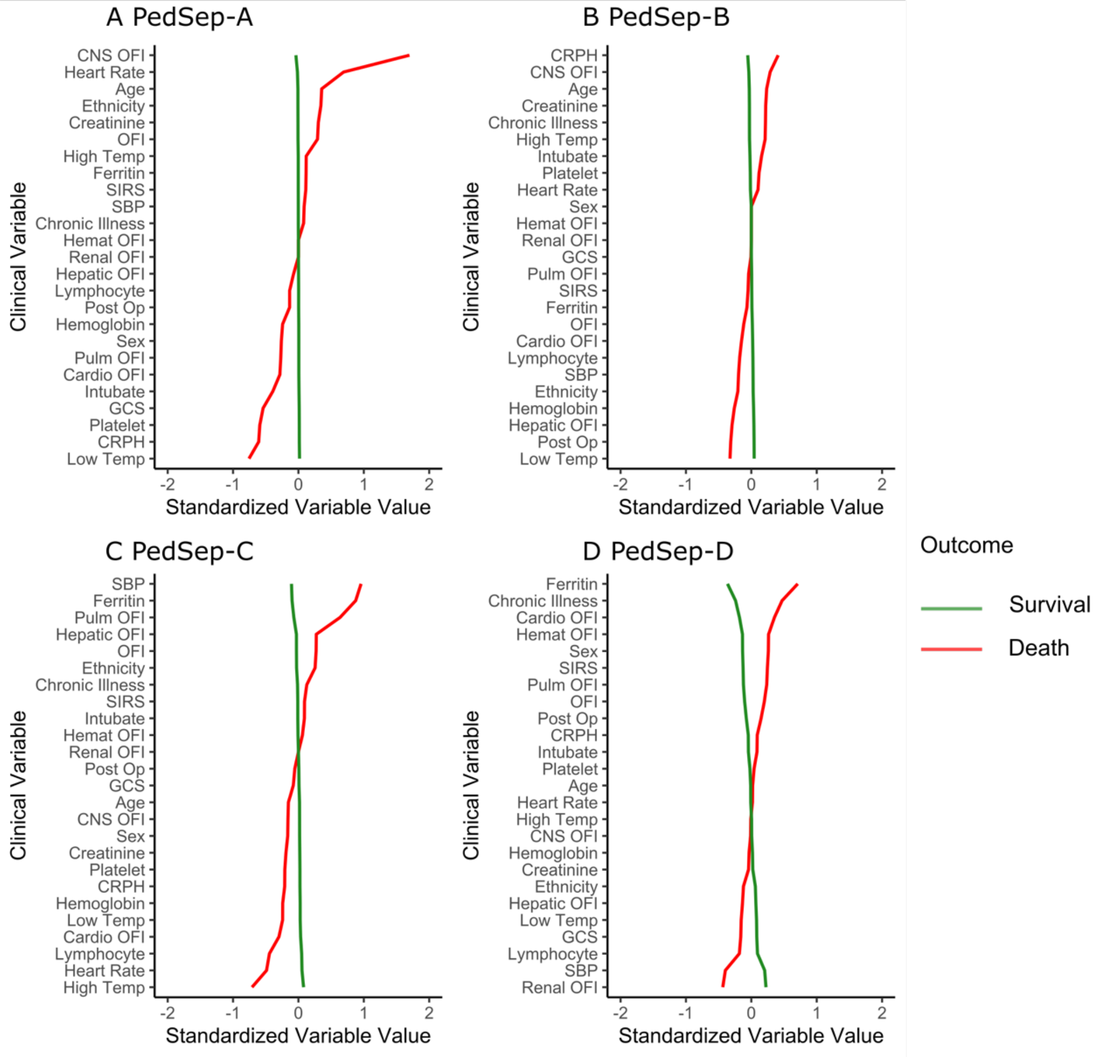


In all panels, the variables are standardized such that all means are scaled to 0 and SDs to 1. A value of 1 for the standardized variable value (x-axis) signifies that the mean value for the phenotype was 1 SD higher than the mean value for both phenotypes shown in the graph as a whole. CNS -central nervous system; CRPH -C-reactive protein; GCS -Glasgow Coma Scale; Hemat -Hematologic; Intubate-Intubation with endotracheal tube; OFI-organ failure index; Post Op-post-surgery; Pulm-pulmonary; Temp-temperature; SBP-systolic blood pressure; Chronic illness –those who are not recorded as previous healthy in table 1 and table 2; Ethnicity –value is higher with more non-Hispanics in group; Sex –value is higher with more males in group;

#### eFigure 12. Comparison of Day 1 Biomarkers That Contribute to Survival in PedSep-A, B, C, and D

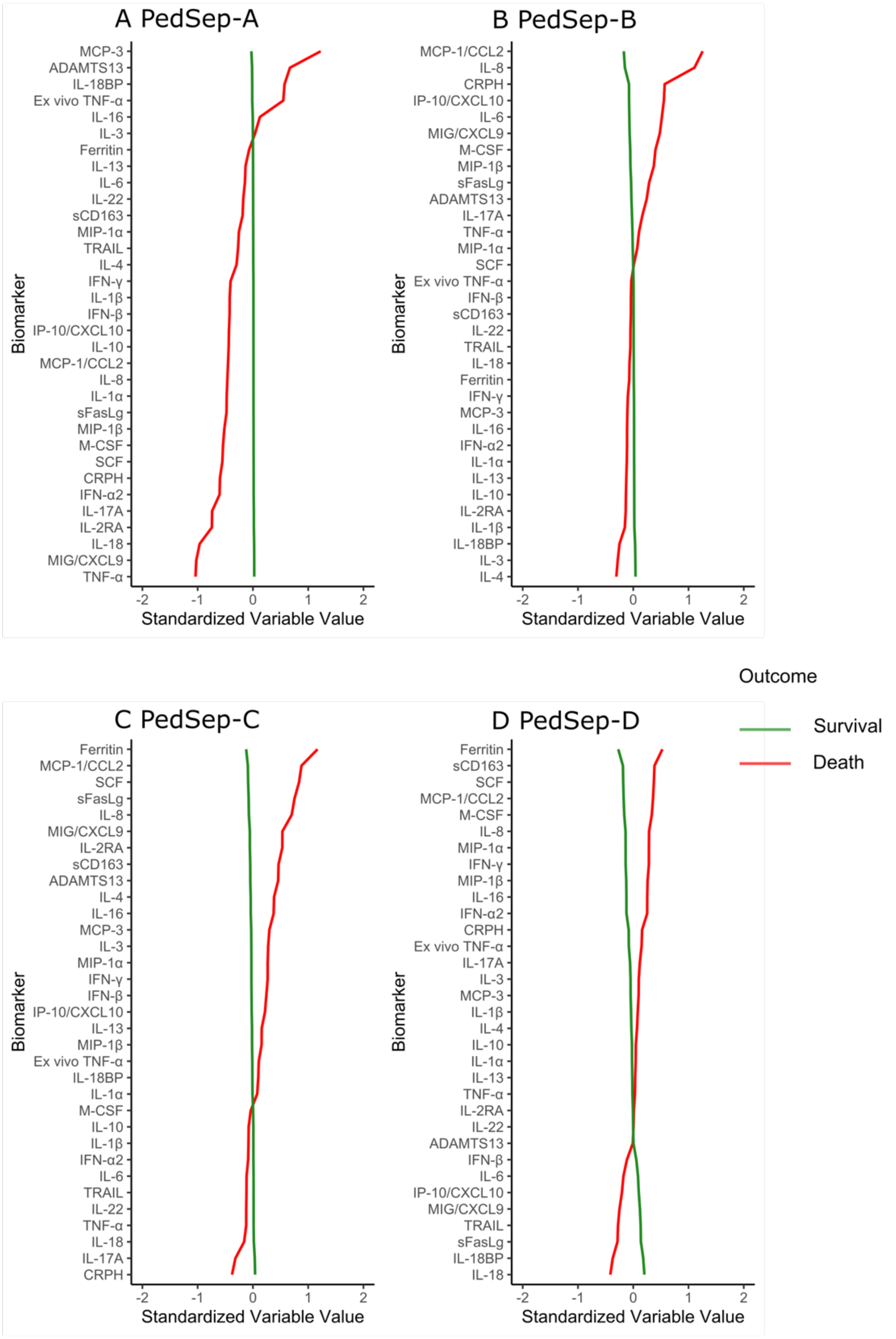


In all panels, the variables are standardized such that all means are scaled to 0 and SDs to 1. A value of 1 for the standardized variable value (x-axis) signifies that the mean value for the phenotype was 1 SD higher than the mean value for both phenotypes shown in the graph as a whole.

#### eFigure 13. t-SNE plot of outcomes (N = 404) across PedSep-A, B, C, and D

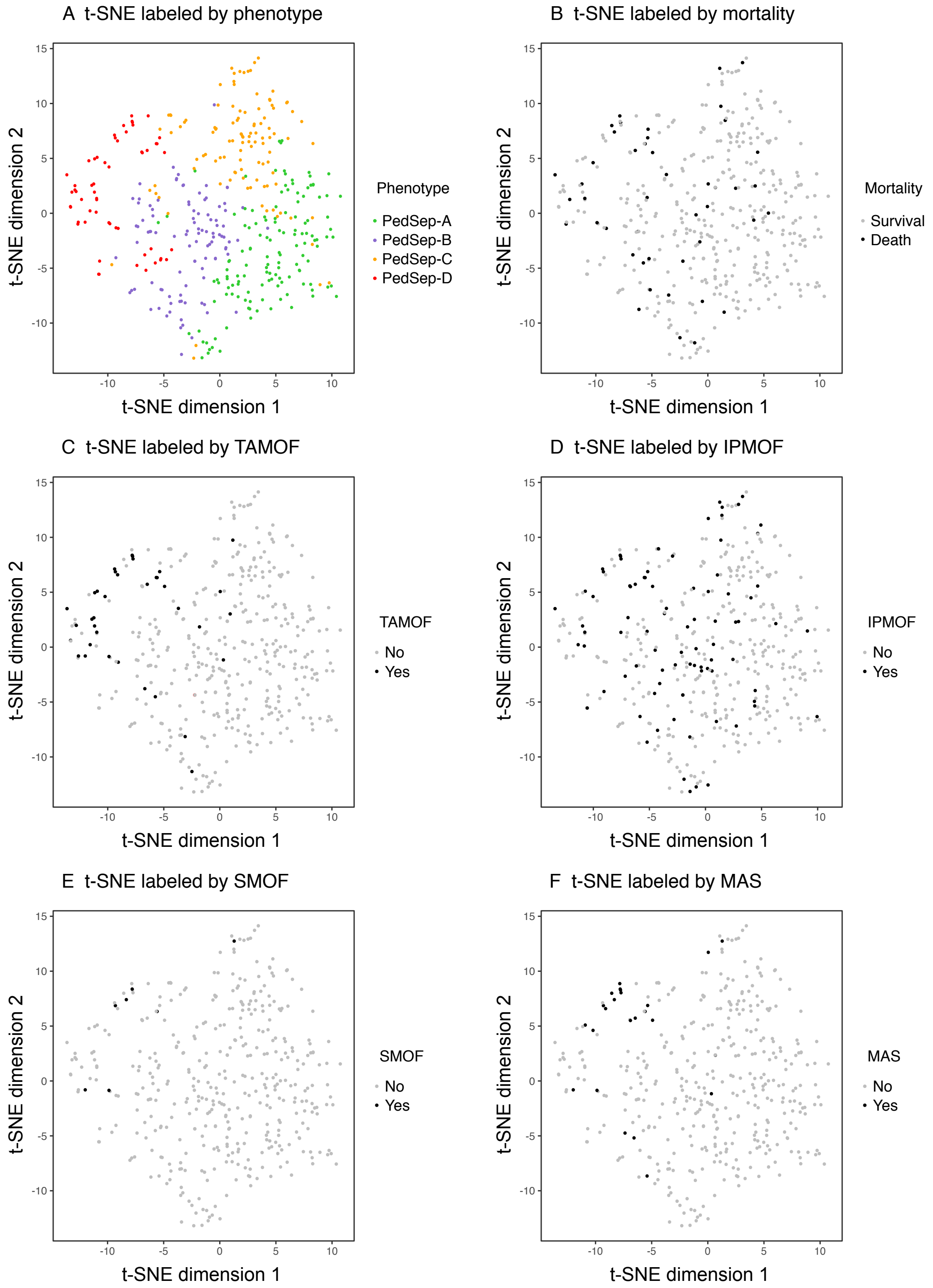


Abbreviations: TAMOF, thrombocytopenia associated multiple organ failure; IPMOF, immunoparalysis associated multiple organ failure; SMOF, sequential liver failure associated multiple organ failure; MAS, macrophage activation syndrome

(A) Visualization of phenotypes using t-distributed stochastic neighbor embedding (t-SNE) technique with phenotype shown in color, (B) mortality shown in color, (B) TAMOF shown in color, (C) IPMOF shown in color, (D) SMOF shown in color, and (E) MAS shown in color

### eFigure 14. Counts of patients (14 therapies)

A Count of treated patients

PedSep-B

| | ANAKINRA | CYCLOSPORINE | DECADRON | ETOPOSIDE | HYDROXYUREA | IMMUNOGLOBULIN G | METHYLPREDNISOLONE | MYCOPHENOLATE | NEUPOGEN | TACROLIMUS | CRRT | ECMO | MechVent | PlasmaExchange |
|---|---|---|---|---|---|---|---|---|---|---|---|---|---|---|
| ANAKINRA | 1 | 0 | 1 | 0 | 0 | 1 | 0 | 0 | 0 | 0 | 0 | 0 | 1 | 0 |
| CYCLOSPORINE | 0 | 0 | 0 | 0 | 0 | 0 | 0 | 0 | 0 | 0 | 0 | 0 | 0 | 0 |
| DECADRON | 1 | 0 | 22 | 0 | 0 | 3 | 7 | 0 | 0 | 0 | 1 | 1 | 22 | 0 |
| ETOPOSIDE | 0 | 0 | 0 | 0 | 0 | 0 | 0 | 0 | 0 | 0 | 0 | 0 | 0 | 0 |
| HYDROXYUREA | 0 | 0 | 0 | 0 | 0 | 0 | 0 | 0 | 0 | 0 | 0 | 0 | 0 | 0 |
| IMMUNOGLOBULIN G | 1 | 0 | 3 | 0 | 0 | 10 | 4 | 0 | 0 | 0 | 1 | 1 | 10 | 0 |
| METHYLPREDNISOLONE | 0 | 0 | 7 | 0 | 0 | 4 | 23 | 0 | 0 | 0 | 2 | 2 | 23 | 0 |
| MYCOPHENOLATE | 0 | 0 | 0 | 0 | 0 | 0 | 0 | 1 | 0 | 1 | 0 | 0 | 0 | 0 |
| NEUPOGEN | 0 | 0 | 0 | 0 | 0 | 0 | 0 | 0 | 2 | 0 | 1 | 0 | 2 | 0 |
| TACROLIMUS | 0 | 0 | 0 | 0 | 0 | 0 | 0 | 1 | 0 | 3 | 0 | 0 | 2 | 0 |
| CRRT | 0 | 0 | 1 | 0 | 0 | 1 | 2 | 0 | 1 | 0 | 5 | 1 | 5 | 0 |
| ECMO | 0 | 0 | 1 | 0 | 0 | 1 | 2 | 0 | 0 | 0 | 1 | 7 | 7 | 1 |
| MechVent | 1 | 0 | 22 | 0 | 0 | 10 | 23 | 0 | 2 | 2 | 5 | 7 | 77 | 2 |
| PlasmaExchange | 0 | 0 | 0 | 0 | 0 | 0 | 0 | 0 | 0 | 0 | 0 | 1 | 2 | 2 |

PedSep-C

| | ANAKINRA | CYCLOSPORINE | DECADRON | ETOPOSIDE | HYDROXYUREA | IMMUNOGLOBULIN G | METHYLPREDNISOLONE | MYCOPHENOLATE | NEUPOGEN | TACROLIMUS | CRRT | ECMO | MechVent | PlasmaExchange |
|---|---|---|---|---|---|---|---|---|---|---|---|---|---|---|
| ANAKINRA | 3 | 1 | 2 | 2 | 0 | 3 | 2 | 1 | 3 | 1 | 1 | 0 | 2 | 0 |
| CYCLOSPORINE | 1 | 1 | 1 | 1 | 0 | 1 | 1 | 1 | 1 | 1 | 1 | 0 | 1 | 0 |
| DECADRON | 2 | 1 | 14 | 2 | 0 | 5 | 5 | 2 | 3 | 2 | 3 | 3 | 13 | 1 |
| ETOPOSIDE | 2 | 1 | 2 | 2 | 0 | 2 | 1 | 1 | 2 | 1 | 1 | 0 | 2 | 0 |
| HYDROXYUREA | 0 | 0 | 0 | 0 | 0 | 0 | 0 | 0 | 0 | 0 | 0 | 0 | 0 | 0 |
| IMMUNOGLOBULIN G | 3 | 1 | 5 | 2 | 0 | 19 | 9 | 3 | 9 | 4 | 6 | 0 | 16 | 3 |
| METHYLPREDNISOLONE | 2 | 1 | 5 | 1 | 0 | 9 | 24 | 4 | 5 | 5 | 3 | 4 | 22 | 3 |
| MYCOPHENOLATE | 1 | 1 | 2 | 1 | 0 | 3 | 4 | 4 | 1 | 3 | 2 | 0 | 4 | 0 |
| NEUPOGEN | 3 | 1 | 3 | 2 | 0 | 9 | 5 | 1 | 12 | 2 | 3 | 0 | 10 | 1 |
| TACROLIMUS | 1 | 1 | 2 | 1 | 0 | 4 | 5 | 3 | 2 | 5 | 2 | 0 | 5 | 0 |
| CRRT | 1 | 1 | 3 | 1 | 0 | 6 | 3 | 2 | 3 | 2 | 7 | 1 | 7 | 2 |
| ECMO | 0 | 0 | 3 | 0 | 0 | 0 | 4 | 0 | 0 | 0 | 1 | 5 | 5 | 1 |
| MechVent | 2 | 1 | 13 | 2 | 0 | 16 | 22 | 4 | 10 | 5 | 7 | 5 | 60 | 4 |
| PlasmaExchange | 0 | 0 | 1 | 0 | 0 | 3 | 3 | 0 | 1 | 0 | 2 | 1 | 4 | 4 |

PedSep-D

| | ANAKINRA | CYCLOSPORINE | DECADRON | ETOPOSIDE | HYDROXYUREA | IMMUNOGLOBULIN G | METHYLPREDNISOLONE | MYCOPHENOLATE | NEUPOGEN | TACROLIMUS | CRRT | ECMO | MechVent | PlasmaExchange |
|---|---|---|---|---|---|---|---|---|---|---|---|---|---|---|
| ANAKINRA | 1 | 1 | 0 | 0 | 1 | 1 | 1 | 0 | 0 | 0 | 1 | 0 | 1 | 1 |
| CYCLOSPORINE | 1 | 1 | 0 | 0 | 1 | 1 | 1 | 0 | 0 | 0 | 1 | 0 | 1 | 1 |
| DECADRON | 0 | 0 | 8 | 1 | 0 | 2 | 3 | 1 | 1 | 0 | 4 | 2 | 7 | 3 |
| ETOPOSIDE | 0 | 0 | 1 | 2 | 0 | 1 | 1 | 0 | 0 | 0 | 1 | 0 | 1 | 2 |
| HYDROXYUREA | 1 | 1 | 0 | 0 | 2 | 1 | 1 | 0 | 1 | 0 | 2 | 0 | 2 | 1 |
| IMMUNOGLOBULIN G | 1 | 1 | 2 | 1 | 1 | 16 | 7 | 0 | 4 | 3 | 10 | 4 | 14 | 3 |
| METHYLPREDNISOLONE | 1 | 1 | 3 | 1 | 1 | 7 | 16 | 3 | 3 | 5 | 11 | 6 | 14 | 3 |
| MYCOPHENOLATE | 0 | 0 | 1 | 0 | 0 | 0 | 3 | 3 | 1 | 2 | 3 | 2 | 3 | 1 |
| NEUPOGEN | 0 | 0 | 1 | 0 | 1 | 4 | 3 | 1 | 9 | 0 | 6 | 3 | 9 | 2 |
| TACROLIMUS | 0 | 0 | 0 | 0 | 0 | 3 | 5 | 2 | 0 | 7 | 7 | 1 | 6 | 1 |
| CRRT | 1 | 1 | 4 | 1 | 2 | 10 | 11 | 3 | 6 | 7 | 34 | 9 | 32 | 9 |
| ECMO | 0 | 0 | 2 | 0 | 0 | 4 | 6 | 2 | 3 | 1 | 9 | 10 | 10 | 3 |
| MechVent | 1 | 1 | 7 | 1 | 2 | 14 | 14 | 3 | 9 | 6 | 32 | 10 | 46 | 10 |
| PlasmaExchange | 1 | 1 | 3 | 2 | 1 | 3 | 3 | 1 | 2 | 1 | 9 | 3 | 10 | 11 |

B Count of survived patients

PedSep-B

| | ANAKINRA | CYCLOSPORINE | DECADRON | ETOPOSIDE | HYDROXYUREA | IMMUNOGLOBULIN G | METHYLPREDNISOLONE | MYCOPHENOLATE | NEUPOGEN | TACROLIMUS | CRRT | ECMO | MechVent | PlasmaExchange |
|---|---|---|---|---|---|---|---|---|---|---|---|---|---|---|
| ANAKINRA | 1 | 0 | 1 | 0 | 0 | 1 | 0 | 0 | 0 | 0 | 0 | 0 | 1 | 0 |
| CYCLOSPORINE | 0 | 0 | 0 | 0 | 0 | 0 | 0 | 0 | 0 | 0 | 0 | 0 | 0 | 0 |
| DECADRON | 1 | 0 | 19 | 0 | 0 | 2 | 6 | 0 | 0 | 0 | 0 | 0 | 19 | 0 |
| ETOPOSIDE | 0 | 0 | 0 | 0 | 0 | 0 | 0 | 0 | 0 | 0 | 0 | 0 | 0 | 0 |
| HYDROXYUREA | 0 | 0 | 0 | 0 | 0 | 0 | 0 | 0 | 0 | 0 | 0 | 0 | 0 | 0 |
| IMMUNOGLOBULIN G | 1 | 0 | 2 | 0 | 0 | 7 | 3 | 0 | 0 | 0 | 0 | 0 | 7 | 0 |
| METHYLPREDNISOLONE | 0 | 0 | 6 | 0 | 0 | 3 | 21 | 0 | 0 | 0 | 1 | 2 | 21 | 0 |
| MYCOPHENOLATE | 0 | 0 | 0 | 0 | 0 | 0 | 0 | 1 | 0 | 1 | 0 | 0 | 0 | 0 |
| NEUPOGEN | 0 | 0 | 0 | 0 | 0 | 0 | 0 | 0 | 1 | 0 | 0 | 0 | 1 | 0 |
| TACROLIMUS | 0 | 0 | 0 | 0 | 0 | 0 | 0 | 1 | 0 | 3 | 0 | 0 | 2 | 0 |
| CRRT | 0 | 0 | 0 | 0 | 0 | 0 | 1 | 0 | 0 | 0 | 2 | 1 | 2 | 0 |
| ECMO | 0 | 0 | 0 | 0 | 0 | 0 | 2 | 0 | 0 | 0 | 1 | 3 | 3 | 0 |
| MechVent | 1 | 0 | 19 | 0 | 0 | 7 | 21 | 0 | 1 | 2 | 2 | 3 | 67 | 1 |
| PlasmaExchange | 0 | 0 | 0 | 0 | 0 | 0 | 0 | 0 | 0 | 0 | 0 | 0 | 1 | 1 |

PedSep-C

| | ANAKINRA | CYCLOSPORINE | DECADRON | ETOPOSIDE | HYDROXYUREA | IMMUNOGLOBULIN G | METHYLPREDNISOLONE | MYCOPHENOLATE | NEUPOGEN | TACROLIMUS | CRRT | ECMO | MechVent | PlasmaExchange |
|---|---|---|---|---|---|---|---|---|---|---|---|---|---|---|
| ANAKINRA | 1 | 0 | 0 | 0 | 0 | 1 | 1 | 0 | 1 | 0 | 0 | 0 | 0 | 0 |
| CYCLOSPORINE | 0 | 0 | 0 | 0 | 0 | 0 | 0 | 0 | 0 | 0 | 0 | 0 | 0 | 0 |
| DECADRON | 0 | 0 | 8 | 0 | 0 | 1 | 1 | 0 | 0 | 0 | 1 | 1 | 7 | 1 |
| ETOPOSIDE | 0 | 0 | 0 | 0 | 0 | 0 | 0 | 0 | 0 | 0 | 0 | 0 | 0 | 0 |
| HYDROXYUREA | 0 | 0 | 0 | 0 | 0 | 0 | 0 | 0 | 0 | 0 | 0 | 0 | 0 | 0 |
| IMMUNOGLOBULIN G | 1 | 0 | 1 | 0 | 0 | 13 | 6 | 1 | 5 | 1 | 4 | 0 | 10 | 3 |
| METHYLPREDNISOLONE | 1 | 0 | 1 | 0 | 0 | 6 | 16 | 1 | 2 | 2 | 2 | 2 | 14 | 3 |
| MYCOPHENOLATE | 0 | 0 | 0 | 0 | 0 | 1 | 1 | 1 | 0 | 1 | 1 | 0 | 1 | 0 |
| NEUPOGEN | 1 | 0 | 0 | 0 | 0 | 5 | 2 | 0 | 7 | 0 | 1 | 0 | 5 | 1 |
| TACROLIMUS | 0 | 0 | 0 | 0 | 0 | 1 | 2 | 1 | 0 | 2 | 1 | 0 | 2 | 0 |
| CRRT | 0 | 0 | 1 | 0 | 0 | 4 | 2 | 1 | 1 | 1 | 5 | 1 | 5 | 2 |
| ECMO | 0 | 0 | 1 | 0 | 0 | 0 | 2 | 0 | 0 | 0 | 1 | 3 | 3 | 1 |
| MechVent | 0 | 0 | 7 | 0 | 0 | 10 | 14 | 1 | 5 | 2 | 5 | 3 | 49 | 4 |
| PlasmaExchange | 0 | 0 | 1 | 0 | 0 | 3 | 3 | 0 | 1 | 0 | 2 | 1 | 4 | 4 |

PedSep-D

| | ANAKINRA | CYCLOSPORINE | DECADRON | ETOPOSIDE | HYDROXYUREA | IMMUNOGLOBULIN G | METHYLPREDNISOLONE | MYCOPHENOLATE | NEUPOGEN | TACROLIMUS | CRRT | ECMO | MechVent | PlasmaExchange |
|---|---|---|---|---|---|---|---|---|---|---|---|---|---|---|
| ANAKINRA | 0 | 0 | 0 | 0 | 0 | 0 | 0 | 0 | 0 | 0 | 0 | 0 | 0 | 0 |
| CYCLOSPORINE | 0 | 0 | 0 | 0 | 0 | 0 | 0 | 0 | 0 | 0 | 0 | 0 | 0 | 0 |
| DECADRON | 0 | 0 | 5 | 1 | 0 | 2 | 1 | 0 | 0 | 0 | 2 | 0 | 4 | 1 |
| ETOPOSIDE | 0 | 0 | 1 | 2 | 0 | 1 | 1 | 0 | 0 | 0 | 1 | 0 | 1 | 2 |
| HYDROXYUREA | 0 | 0 | 0 | 0 | 0 | 0 | 0 | 0 | 0 | 0 | 0 | 0 | 0 | 0 |
| IMMUNOGLOBULIN G | 0 | 0 | 2 | 1 | 0 | 9 | 6 | 0 | 3 | 2 | 5 | 3 | 8 | 2 |
| METHYLPREDNISOLONE | 0 | 0 | 1 | 1 | 0 | 6 | 11 | 1 | 2 | 4 | 6 | 2 | 9 | 1 |
| MYCOPHENOLATE | 0 | 0 | 0 | 0 | 0 | 0 | 1 | 1 | 0 | 1 | 1 | 0 | 1 | 0 |
| NEUPOGEN | 0 | 0 | 0 | 0 | 0 | 3 | 2 | 0 | 5 | 0 | 2 | 2 | 5 | 1 |
| TACROLIMUS | 0 | 0 | 0 | 0 | 0 | 2 | 4 | 1 | 0 | 5 | 5 | 0 | 5 | 1 |
| CRRT | 0 | 0 | 2 | 1 | 0 | 5 | 6 | 1 | 2 | 5 | 20 | 3 | 19 | 6 |
| ECMO | 0 | 0 | 0 | 0 | 0 | 3 | 2 | 0 | 2 | 0 | 3 | 4 | 4 | 2 |
| MechVent | 0 | 0 | 4 | 1 | 0 | 8 | 9 | 1 | 5 | 5 | 19 | 4 | 29 | 6 |
| PlasmaExchange | 0 | 0 | 1 | 2 | 0 | 2 | 1 | 0 | 1 | 1 | 6 | 2 | 6 | 7 |

This heatmap shows the count of treated patients (A) and survivors among treated patients (B) in PedSep-B, C, and D. Therapies not associated with outcomes in univariable analysis (eTable 10) were not included. Drugs and Organ Support treatments are sorted in alphabetical order.
